# The preliminary safety and immunogenicity results of a randomized, double-blind, placebo-controlled Phase I trial for a recombinant two-component subunit SARS-CoV-2 vaccine ReCOV

**DOI:** 10.1101/2022.05.11.22274932

**Authors:** Chris Wynne, Paul Hamilton, Jingxin Li, Chen Mo, Jiaping Yu, Wenrong Yao, Zijing Yue, Xi Zhang, Jianhui Zhang, Kunxue Hong, Jianping Chen, Yong Liu, Fengcai Zhu

## Abstract

**Background:** The ReCOV is a recombinant trimeric two-component SARS-CoV-2 subunit vaccine adjuvanted with BFA03. We report the preliminary safety and immunogenicity results for the ReCOV.

**Methods:** This first in human, randomized, double-blind, placebo-controlled phase I study, was conducted at 2 study sites in New Zealand. Subjects were stratified into two age cohorts (18-55 years and 56-80 years old) and then randomly assigned in a 4:1 ratio to receive two 0.5 mL intramuscular doses of the ReCOV vaccine (20µg or 40µg, adjuvanted with BFA03 in each) or placebo, 21 days apart. The primary endpoints were incidence of solicited local and systemic adverse events (AEs) and unsolicited AEs after each dose; incidence of serious adverse events (SAEs) up to 30 days after the second dose; changes in clinical laboratory tests from baseline up to 7 days after each dose; and changes in vital signs from baseline up to 30 days after the second dose. The key secondary endpoints for immunogenicity were neutralizing antibody titers against SARS-CoV-2, S1 receptor binding domain (RBD) and N-terminal domain (NTD) IgG titers post-vaccination. The T cell-specific immune response elicited by ReCOV were also evaluated. The trial was registered with ClinicalTrials.gov (NCT04818801).

**Findings:** One hundred participants (50 for each age group) were randomized. The incidence of solicited local AEs in 20μg ReCOV, 40μg ReCOV, and pooled placebo group among younger adults were 60.0%, 70.0%, and 10.0%, respectively, while among older adults were 55.0%, 84.2%, and 10.0%, respectively. The incidence of solicited systemic AEs in 20μg ReCOV, 40μg ReCOV, and pooled placebo group among younger adults were 60.0%, 60.0%, and 30.0%, respectively, while among older adults were 50.0%, 52.6%, and 50.0%, respectively. All solicited AEs and unsolicited AEs were mild. No vaccination-related SAE, adverse events of special interest, and AE leading to early discontinuation were reported.

ReCOV elicited SARS-CoV-2 neutralizing antibody after the first vaccination, which were increased further after the second vaccination irrespective of dose and age groups. The neutralizing antibody against wild-type SARS-CoV-2 peaked at 14 days post the second vaccination in both 20µg and 40µg ReCOV groups, with GMT of 1643.17 IU/mL and 1289.21 IU/mL among younger adults, and 1122.32 IU/mL and 680.31 IU/mL among older adults, respectively. Similarly, both anti-RBD and anti-NTD specific IgG were elicited after the first vaccination, and peaked at 14 days after the second vaccination. T helper 1 biased cellular responses were observed after ReCOV vaccinations.

**Interpretation:** Both 20 and 40µg ReCOV showed good safety profiles and elicited strong immune responses in the younger and the older adults. The results of this study support the accelerated development of ReCOV.

**Funding:** Jiangsu Recbio Technology Co., Ltd.

## Introduction

During the global pandemic, new coronavirus disease 2019 (COVID-19) caused by severe acute respiratory syndrome coronavirus 2 (SARS-CoV-2) has resulted in considerable illness and death around the world. Up to March 16, 2022, a total of 460,280,168 confirmed cases of COVID-19 and 6,050,018 deaths have been reported in 230 countries or regions around the world.^1^

In response to this worldwide pandemic situation, the urgent needs for safe and effective vaccines to mitigate the global spread of SARS-CoV-2 has been prompted. Up to the time of writing, ten vaccines have been validated for WHO emergency use listing since late 2020, including several recombinant protein vaccines, i.e. the NVX-CoV2372 trimeric nanoparticle produced by Novavax, the S-Trimer vaccine developed by Clover Biopharmaceuticals, both have proved their good immunogenicity and protective efficacy in clinical trials.^2,3^ Additionally, more than 153 candidate COVID-19 vaccines are currently undergoing investigation in clinical trials, and the majority (around 34%) belongs to protein subunit vaccines.^4^ The spike protein is the key to the invasion of cells by SARS-CoV-2 and exists on the surface of the virus membrane in the form of trimers, which is the primary source of vaccine antigens.^5^

ReCOV was expressed in CHO cells, containing the 14-541 amino acid sequence (NTD and RBD domains) of the spike protein and is fused with the foldon of T4 bacteriophage at the C-terminus to form a trimerized protein containing NTD-RBD foldon. Pre-clinical proof-of-concept study showed that the combination of NTD and RBD was superior to either RBD or NTD alone in eliciting neutralizing activity.^6^ The ReCOV is adjuvanted by BFA03 (AS03-like squalene adjuvant), which is an oil-in-water emulsion containing two immunostimulants (squalene, α-tocopherol). Two-dose immunizations of ReCOV, with an interval of 21 days, elicited potent high titers of virus-specific binding and neutralizing antibody responses to SARS-CoV-2, provided complete protection against challenge with SARS-CoV-2 in hACE2 transgenic mice and rhesus macaques.^7^

Here, we reported the preliminary safety and immunogenicity results of ReCOV from a first in human phase I study conducted in New Zealand.

## Methods

### Study design and subjects

This is a randomized, double-blind, placebo-controlled phase I study, conducted at 2 study sites in New Zealand. Healthy subjects aged between 18-55 years or 56-80 years were recruited. Subjects who had history of COVID-19 or confirmed SARS-CoV-2 infection; positive test for COVID-19 at screening by reverse transcriptase polymerase chain reaction, or serological test for SARS-CoV-2 immunoglobulin (Ig)M and/or IgG antibodies; received any prior investigational or approved vaccine against a coronavirus at any time; vaccination of licensed inactivated vaccines within 14 days or licensed live or attenuated vaccines within 30 days prior to enrollment in this study; and pregnant women were excluded.

This study was performed in compliance with International Council for Harmonization Good Clinical Practice, and the ethical principles of the Declaration of Helsinki. Prior to initiation of the study, the study protocol, and informed consent form were reviewed and approved by the Independent Ethics Committees (ie, Health and Disability Ethics Committee). Informed consent was obtained from each subject before any study procedures were performed.

### Randomization and masking

The first 2 sentinel subjects in each cohort were randomized at a 1:1 ratio to receive ReCOV or placebo, and the overall randomization for each cohort was at a ratio of 4:1 to receive ReCOV or placebo. Randomization was performed via the electronic data capture system (Viedoc™). Except from prespecified unblinded individuals, all the other investigator, site staff, sponsor, laboratory staff, and study subjects were blinded to the treatment allocation. Vaccine dose preparation and administration were performed by the unblinded pharmacy personnel who did not participate in any other process of the study. Each subject was individually unblinded to their treatment allocation after completing the visit at Day 52 (30 days after the second dose) so as to allow the placebo recipients to withdraw the study and to receive an authorized/licensed (national rollout) COVID-19 vaccine.

### Procedure

The study progressed in a sequential manner, i.e., from low-dose (20 μg) to high-dose (40 μg) groups, and from younger adults to older adults (appendix **p1**). A safety monitoring committee (SMC), composed of representatives of investigators and sponsors, was established to review the safety data before the enrollment of the next cohort. Younger adults in the low-dose group (Cohort 1) were enrolled and administrated with the first dose, then the younger adults in the high-dose group (Cohort 2) and the older adults in the low-dose group (Cohort 3) were enrolled and vaccinated simultaneously after a review of safety data through to 7 days following the first dose of Cohort 1 by SMC. The older adults in the high-dose group (Cohort 4) were enrolled after the safety data through to 7 days following the first dose of Cohort 3 were assessed and reviewed by the SMC, as well as cumulative data from all previously vaccinated cohorts. In each cohort, subjects received 2 intramuscular doses of ReCOV or placebo 21 days apart. Two sentinel subjects in each cohort were vaccinated first and followed for safety at least 48 hours after the first dose, before the enrollment of the rest subjects of the cohort.

Subjects were observed for 30 min (6 hours for sentinel subjects) at site after each vaccination for assessment of immediate adverse reactions, and were instructed to monitor any adverse events in the eDiary outside sites daily. Solicited local AEs (injection site pain, redness, and swelling), solicited systemic AEs (fatigue, fever, gastrointestinal symptoms (nausea, vomiting, diarrhea), headache, and myalgia (muscle pain)) and body temperature were recorded up to 7 days after each vaccination. Unsolicited adverse events were recorded after each dose, up to 30 days after the second dose, SAEs and adverse events of special interest (AESIs) were recorded until the date of the analysis and will be documented during the whole follow-up of the study. Potential immune-mediated diseases, COVID-19 vaccine-specific AEs and COVID-19-specific AEs based on the Coalition for Epidemic Preparedness Innovations Safety Platform for Emergency vaccines Project guidelines were defined as AESIs.^8^ All AEs were graded for severity as mild (Grade 1), moderate (Grade 2), severe (Grade 3), or potentially life-threatening (Grade 4) using Food and Drug Administration Toxicity Grading Scale for Healthy Adult and Adolescent Volunteers Enrolled in Preventive Vaccine Clinical Trials.^9^ Investigators judged the severity gradings and assessed causality as either related or unrelated. Any subject who tested positive for SARS-CoV-2 infection and/or has been confirmed COVID-19 disease (using the WHO COVID-19 case definitions^10^) before completing the two-dose regimen, were withdrawn from the second dose vaccination.

Blood samples for clinical laboratory evaluations (hematology, coagulation, clinical chemistry, urinalysis, and viral serology screening) were collected at Day 1 (before the first dose), at 7 days post the first and second vaccinating, respectively. The samples were analyzed at the study site’s laboratory.

### Outcomes

The primary endpoints were incidence of solicited local and systemic AEs up to 7 days after each dose; incidence of unsolicited AEs after each dose up to 30 days after the second dose; incidence of SAEs up to 30 days after the second dose; changes in clinical laboratory tests from baseline up to 7 days after each dose; and changes in vital signs from baseline up to 30 days after the second dose. The secondary safety endpoints include incidence of AEs, SAEs, AESIs, changes in vital signs from baseline up to 12 months after the second dose. S-RBD and S-NTD IgG binding antibody titers and neutralizing antibody titers against wild-type SAS-CoV-2, as well as the specific T cell responses were evaluated as the secondary endpoints.

We evaluated the immunogenicity elicited by ReCOV vaccine in terms of S-RBD and S-NTD IgG binding antibody titers and neutralizing antibody titers against wild-type SAS-CoV-2. Blood samples were collected at Day 1 (before the first dose), Day 22 (before the second dose), Day 36 (14 days after the second dose), and Day 52 (30 days after the second dose), respectively. Detection of the SARS-CoV-2 neutralizing antibodies was performed using a validated laboratory technique of SARS-CoV-2 microneutralizing assay (cytopathic effect) by the central laboratory 360Biolabs (Melbourne VIC, Australia), with a wild-type strain SARS-CoV-2 hCoV-19/Australia/VIC01/2020 (GenBank MT007544.1) passaged in Vero E6 cells. Sequence analysis of the spike protein showed high sequence homology between SARS-CoV-2 hCoV-19/Australia/VIC01/2020 and the Wuhan strain with only one nucleotide difference in the spike protein (S247R). In addition, 360biolabs used the first WHO International Standard (Catalog #20/136) to calibrate its in-house standards and subsequently converted its assay results to International Units (IU/mL). Neutralizing antibodies against Delta variant were also detected as a post-hoc measurement performed by 360Biolabs using SARS-CoV-2-hCoV-19/Australia/VIC18440/2021 (Delta B.1.617.2 lineage), which was from the Peter Doherty Institute for Infection and Immunity (Melbourne, Australia). S-RBD and S-NTD specific IgG binding antibodies in serum were measured using a validated laboratory technique of V-Plex SARS-CoV-2 Panel 1 (IgG) Multiplex ELISA (MSD Cat # MESOK15359U-4) by 360Biolabs.

Peripheral blood mononuclear cells were isolated from blood samples of all the subjects at baseline, Day 36 and Day 52, respectively, to evaluate the T cell-specific immune responses after vaccination. The secretion of interferon (IFN)-γ, IL-2, IL-4 and IL-5 were measured after stimulating peripheral blood mononuclear cells with SARS-CoV-2 Spike glycoprotein pool. The CD4^+^/CD8^+^ T cells expressing Th1 cytokines (INF-γ/IL-2) and Th2 cytokines (IL-4/IL-5) were assessed using flow cytometry.

### Statistical analysis

As a first in human study, the sample size estimation and power calculation were not pre-specified in the protocol, and the number of proposed subjects was considered sufficient to provide a descriptive summary of the safety and immunogenicity of two different dose levels of ReCOV in two age groups. Data were presented by age groups (younger adults and older adults) and by treatment group. Subjects were grouped and analyzed according to the vaccination they received (20µg ReCOV group, and 40µg ReCOV group, and the pooled placebo group).

Continuous variables were summarized using mean or geometric mean with standard deviation, or median with interquartile range. For categorical variables, frequency and percentage (%) of subjects were presented. The default significance level was 5%, all confidence intervals (CIs) reported were 95% CIs, and all statistical tests were two-sided, unless otherwise specified in the description of the analyses. Demographic and baseline clinical characteristics analysis were performed in all randomized subjects. Safety analysis cohort involved all subjects who randomized and received at least one dose vaccination. The immunogenicity analysis included all subjects in the safety analysis cohort who had at least one quantifiable immunogenicity sample after vaccination. Antibody titers were log-transferred before using for calculation of GMTs and geometric mean fold rises (GMFRs). The antibody titers reported as below the lower limit of quantification were replaced by half of the limit value. While, titers that were greater than the upper limit of quantification were converted to the upper limit. The seroconversion rate (SCR) was defined as the proportion of subjects with at least 4-fold increase in post vaccination antibody titers over baseline.

All analyses were conducted using SAS Version 9.4 or higher (SAS Institute).

The data supporting the statistical analysis depended on the primary analysis data of ReCOV phase I study, referring to all subjects’ data till Day 52 visit.

## Results

Between June 18, 2021 and October 21, 2021, 136 individuals, including 70 aged between 18-55 years, and 66 aged between 56-80 years were recruited and screened. A total of 100 eligible subjects were stratified by age with 50 in each age group, and then randomly assigned either to receive ReCOV (20, 40µg) or placebo (figure 1). Two older adults withdrew, with one subject in Cohort 3 withdrawing from the consent before receiving any vaccination and the other in Cohort 4 withdrawing from the second dose of ReCOV at 40µg due to protocol deviation (positive for Hepatitis B core antibodies). The demographic and baseline clinical characteristics of the subjects (table 1) were comparable across the treatment groups, in both younger and older adults with mean age ranging from 27.8 to 35.5 years and from 60.1 to 61.8 years, and mean body mass index (BMI) ranging from 24.3 to 25.7 kg/m^2^ and from 25.1 to 26.8 kg/m^2^, respectively. The distribution of male and female across the treatment groups in both age groups were similar. In the study population, majority of subjects were White (55.0%) and Asian (35.0%). The ethnicity of subjects was mostly “Not Hispanic or Latino” (75.0%).

**Table 1:**
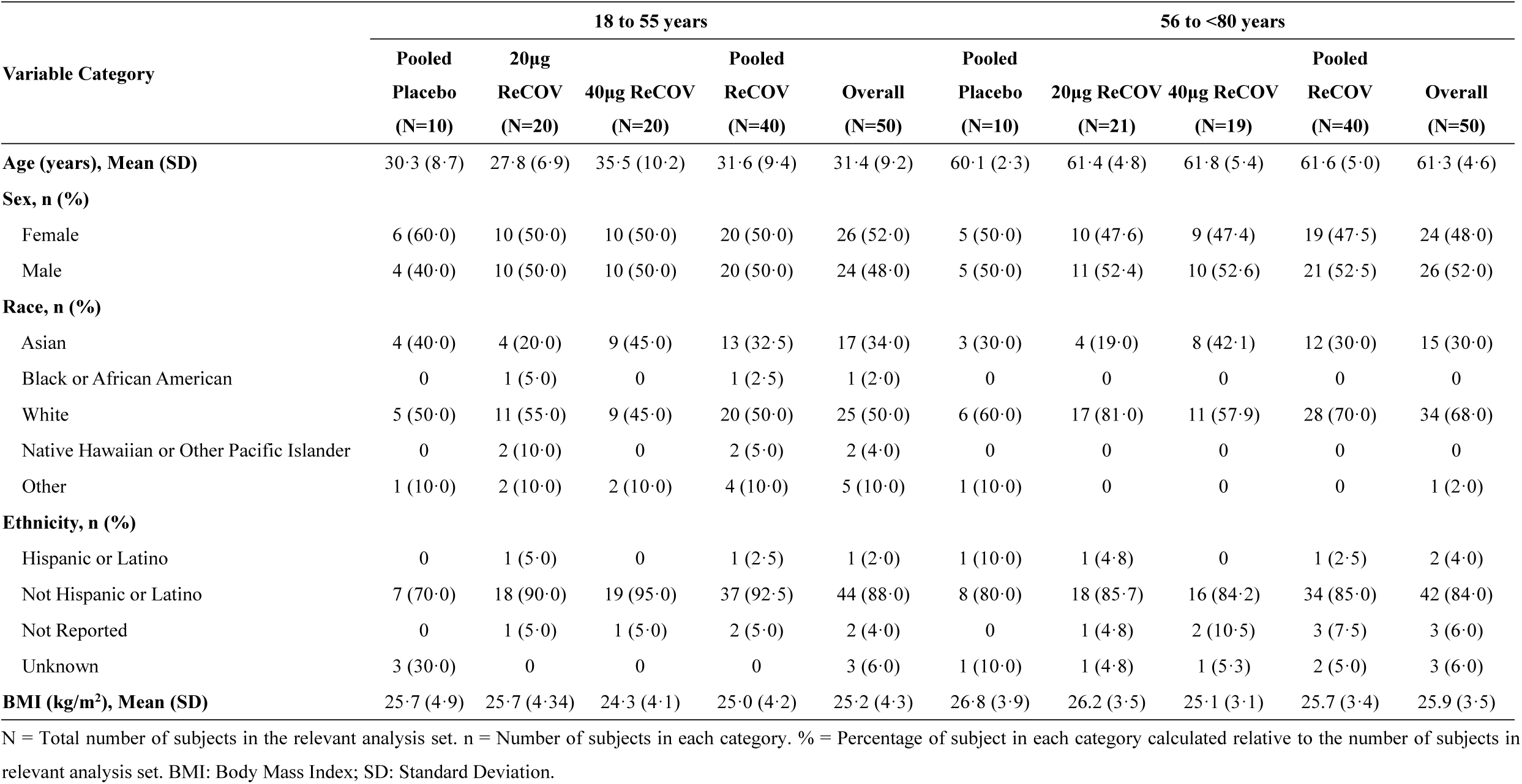
Subjects demographic characteristics (full analysis set) per age group.

**Figure 1:**
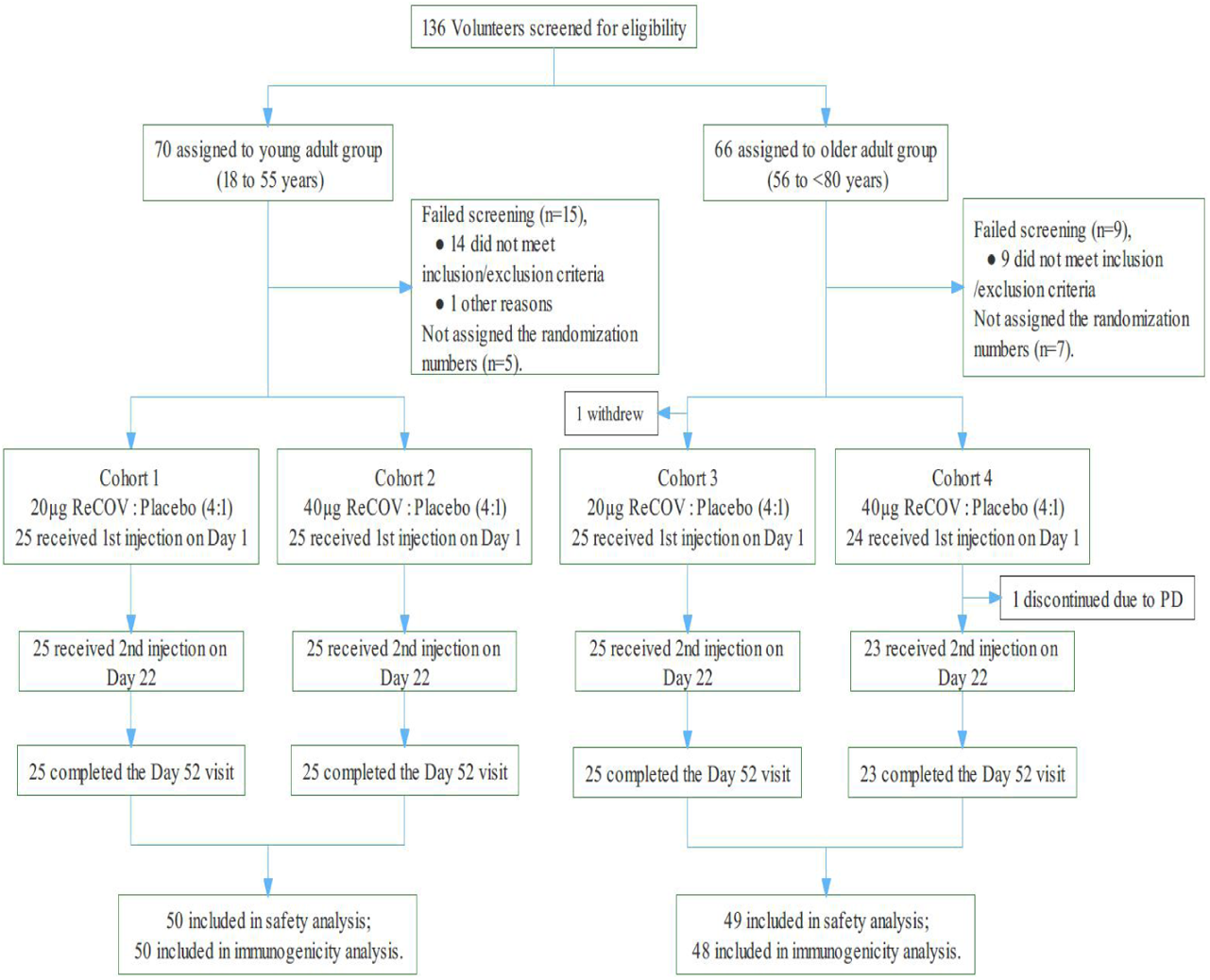
Trial flow diagram. SMC: Safety Monitoring Committee. Cohort 1 will be enrolled and dosed first. Cohorts 2 and 3 will be enrolled and dosed in parallel, after a review of safety data through to 7 days following the first dose from Cohort 1 by the SMC. Cohort 4 will be dosed after a review of safety data through to 7 days following the first dose from Cohort 3, as well as cumulative data from all previously completed cohorts, if available.

The occurrence of solicited local and systemic AEs were similar between younger and older adult subjects who received at least one dose (figure 2). In addition, incidence rate of solicited systemic AEs was similar in subjects receiving 20μg and 40μg ReCOV in both age groups. In both age groups, incidence of solicited local and systemic AEs after receiving the second dosing of 20µg or 40µg ReCOV tended to be higher than that after receiving the first dosing.

**Figure 2:**
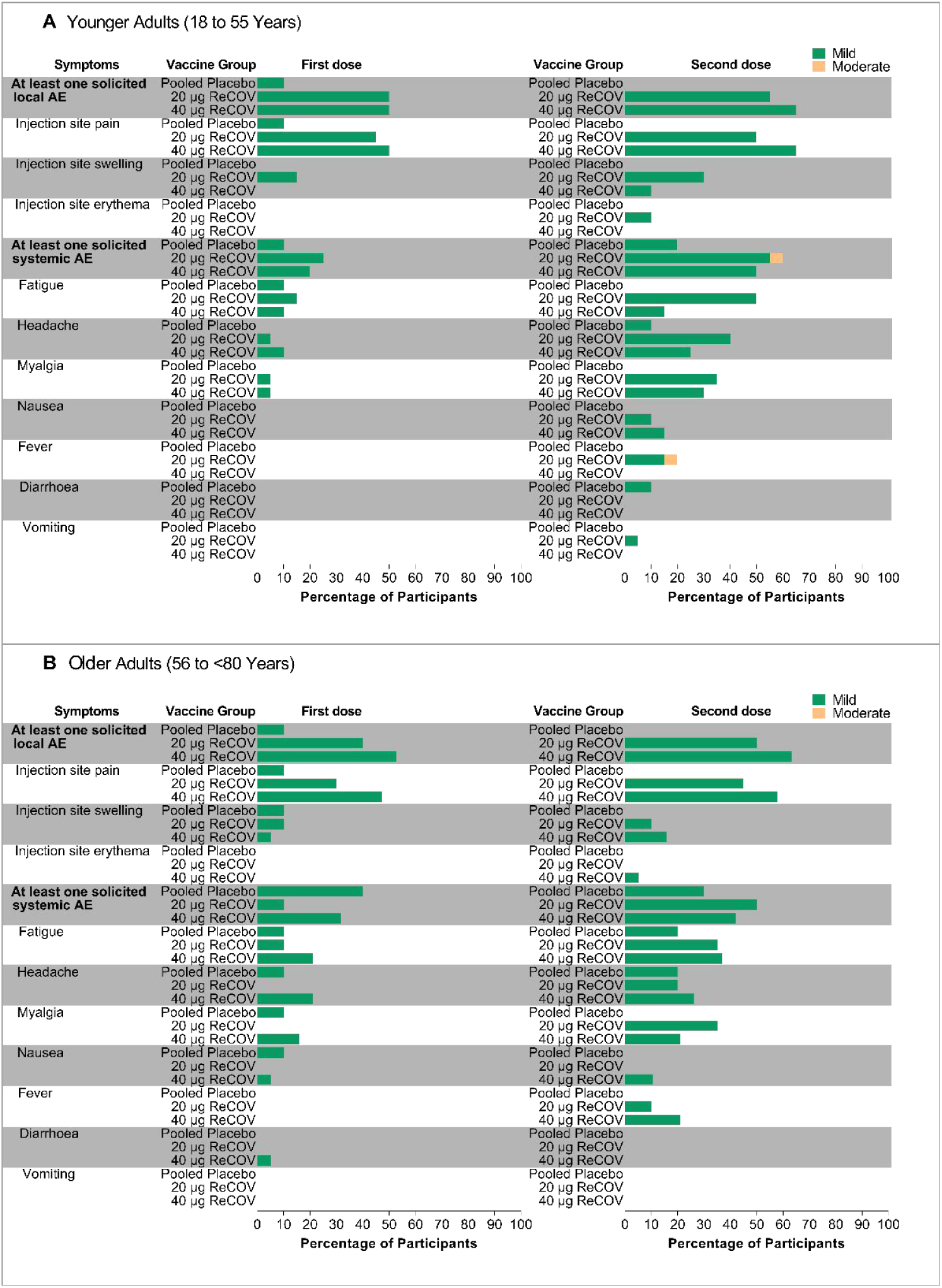
Frequency of solicited local and systemic AEs after the first or second dose in subjects with two dose levels of ReCOV among younger adults (Panel A) and older adults (Panel B)

Among subjects receiving 20μg ReCOV, 40μg ReCOV, or placebo, the incidence of solicited local AEs were 60.0% (12/20), 70.0% (14/20), and 10.0% (1/10) in younger adults, and were 55.0% (11/20), 84.2% (16/19), and 10.0% (1/10) in older adults, respectively. Among subjects receiving 20µg and 40µg ReCOV vaccination, the incidence of solicited local AEs in younger adults were 50% (10/20) and 50% (10/20) after the first dosing, and were 55% (11/20) and 65% (13/20) after the second dosing, while in older adults were 40% (8/20) and 52.6% (10/19) after the first dosing, and were 50% (10/20) and 63.2% (12/19) after the second dosing, respectively (table S1, table S2). In both age groups, the most common ReCOV-related solicited local AE was injection-site pain, with the incidences of 30.0% to 65.0% in 20μg and 40μg ReCOV groups (figure 2). All solicited local AEs after each vaccination were mild, with mean duration of 2.3-3.8 days and 2.8-12.0 days in the younger and older adult group who received ReCOV vaccination, respectively (table S3).

Among subjects receiving 20μg ReCOV, 40μg ReCOV, or placebo, incidence of solicited systemic AEs were 60.0% (12/20), 60.0% (12/20), and 30.0% (3/10) in younger adults, while were 50.0% (10/20), 52.6% (10/19), and 50.0% (5/10) in older adults, respectively. Among subjects receiving 20µg and 40µg ReCOV vaccination, the incidence of solicited systemic AEs in younger adults were 25% (5/20) and 20% (4/20) after the first dosing, and were 60% (12/20) and 50% (10/20) after the second dosing, while in older adults were 10% (2/20) and 31.6% (6/19) after the first dosing, and were 50% (10/20) and 42.1% (8/19) after the second dosing, respectively (table S1, table S4). In both age groups, the common ReCOV-related solicited systemic AEs (≥10%) in either 20μg or 40μg ReCOV group included fatigue, headache, myalgia, pyrexia and nausea (figure 2). Except for one younger adult in 20μg ReCOV group developed moderate (Grade 2) pyrexia, all other solicited systemic AEs were mild, with mean duration of 2.0-4.3 days and 1.8-12.8 days in younger and older adults who received ReCOV vaccination, respectively (table S5).

Among subjects receiving 20μg ReCOV, 40μg ReCOV, or placebo, the incidence of unsolicited AEs was 70.0% (14/20), 60.0% (12/20), and 20.0% (2/10) in younger adults, while were 55.0% (11/20), 63.2% (12/19), and 70.0% (7/10) in older adults, respectively (table S1). Majority unsolicited AEs were mild, except for six moderate (Grade 2) AEs occurred in 5 subjects (3 in ReCOV group, 2 in placebo group). Three moderate AEs occurred in ReCOV group included 1 toothache (unrelated, in a younger adult receiving 20μg), 1 influenza like illness (related, in a younger adult receiving 40μg), and 1 injection site rash (related, in a younger adult receiving 40μg), respectively. Another 3 moderate AEs in placebo group included rhinalgia, toothache, and rash, respectively (table S6).

One SAE occurred in an older adult receiving 20µg ReCOV, which was a tibia fracture and considered as unrelated to the vaccination. No subject experienced related SAE, AESI, AEs ≥ Grade 3, or AE leading to early discontinuation. No clinically significant changes in vital signs and laboratory parameters were identified. SARS-CoV-2 neutralizing antibodies against wild-type strain could be elicited in majority vaccinated subjects at Day 22 after the first vaccination with 20μg or 40μg ReCOV, with the SCR in 20μg and 40μg ReCOV group of 95.0% and 100.0% in younger adults, and of 70.0% and 94.4% in older adults, respectively. At 14 days post the second vaccination (Day 36), the SCRs reached to 100% in all ReCOV groups, irrespective of dose levels and subject ages. No subject in placebo groups showed seropositive at all time points (table S7).

The GMTs of neutralizing antibodies reached peak level at 14 days post the second vaccination (Day 36) and remained at high level at 30 days post the second vaccination (Day 52). Among younger adults in 20μg and 40μg ReCOV groups, the GMTs of neutralizing antibody were 117.97 IU/mL and 135.51 IU/mL, 1643.17 IU/mL and 1289.21 IU/mL, and 1047.16 IU/mL and 740.46 IU/mL at Day 22, 36 and 52, respectively, and the corresponding GMFRs at each time point were 19.03 and 21.86, 265.03 and 207.94, and 168.90 and 119.43, respectively. In older adults, there were approximately 19%-50% reduction in neutralizing antibodies at each time point compared with that in younger adults, however the GMTs in both 20μg or 40μg groups were still high, with 58.99 IU/mL and 78.74 IU/mL, 1122.32 IU/mL and 680.31 IU/mL, and 850.56 IU/mL and 561.16 IU/mL at Day 22, 36 and 52, respectively (figure 3).

**Figure 3:**
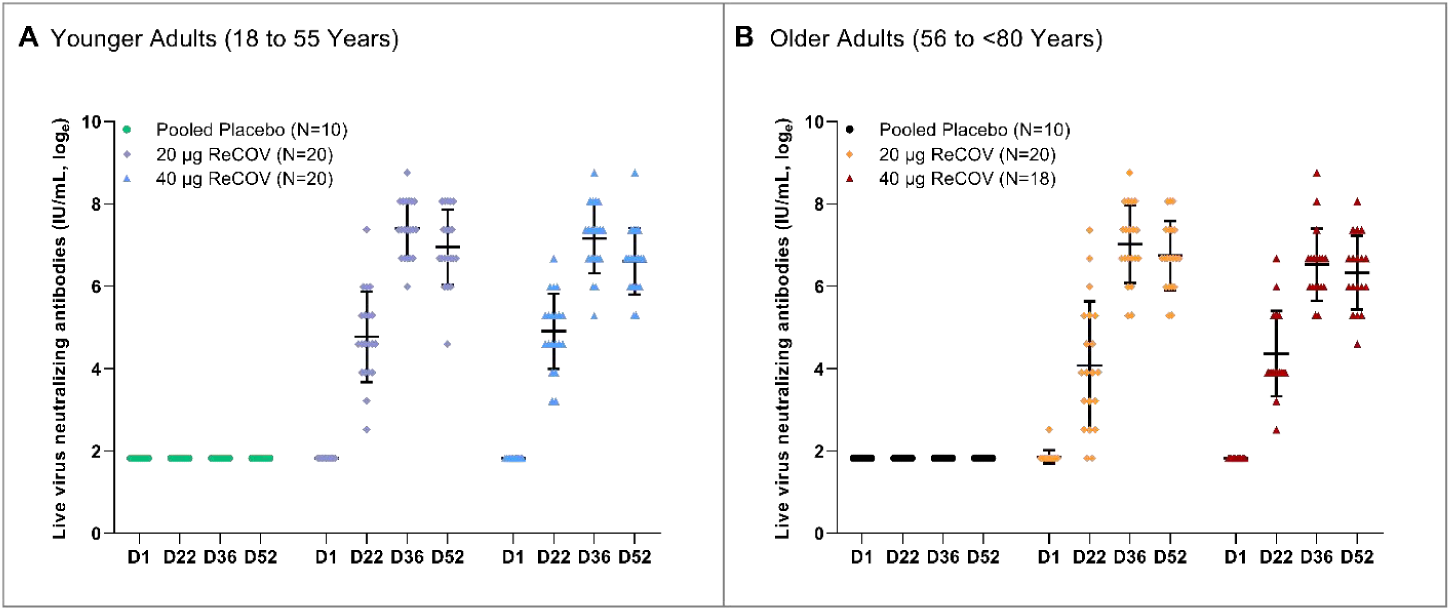
Neutralizing antibody responses (GMT) after the first and second doses by age group. The titers (GMT) of Neutralizing antibody against hCoV-19 Australia. The neutralizing antibody titers were reported at Day 1 (D1), Day 22 (D22), Day 36 (D36) and Day 52 (D52) of vaccinated groups (pooled placebo, 20µg ReCOV, and 40µg ReCOV) for younger adults (Panel A) and older adults (Panel B), respectively by natural base of log transformation. The LLOQ and ULOQ are based on the sample dilution factor which can range from 1 in 100 (minimum) to 1 in 40,000 to ensure that the sample falls within the standard curve.

As post-hoc analyses, the neutralizing antibodies against Delta variant were tested in ReCOV groups at Day 36, 14 days post the second vaccination, which showed approximately 5-11 times reduction in the GMTs, compared to the neutralization activity against wild-type SARS-CoV-2 (table S8). However, the SCRs of Delta variant neutralizing antibodies were 100% at Day 36 in younger adults receiving 20μg and 40μg ReCOV, and were 90% and 88.9% in older adults receiving 20μg and 40μg ReCOV, respectively (table S7).

Both S-RBD and S-NTD specific IgGs were at a low level at baseline. After the first vaccination, the SCRs reached 100% in all ReCOV groups, irrespective of dose levels and subject ages, while in the placebo groups, only one older adult showed seropositive for both antigen-specific antibodies. After the second vaccination, all subjects in placebo group kept seronegative for both antibodies at all time points (table S9).

Similar to neutralizing antibodies, both S-RBD and S-NTD specific IgGs reached peak titers at 14 days post the second vaccination (Day 36), and remained at high levels at Day 52, irrespective of dose levels and subject ages. Among younger adults receiving 20μg or 40μg ReCOV, the peak GMTs of S-RBD specific IgG were 278596.07 AU/mL and 271838.24 AU/mL, and of S-NTD specific IgG were 11232.42 AU/mL and 11266.48 AU/mL, respectively. Among older adults, the peak GMTs were approximately 1.5 times lower compared to younger adults, however the antibody levels were still high, the peak GMTs for S-RBD specific IgG were 185337.57 AU/mL and 203829.72 AU/mL, the peak GMTs for S-NTD specific IgG were 9422.77 AU/mL and 6705.86 AU/mL, in 20μg and 40μg ReCOV group, respectively (figure 4).

**Figure 4:**
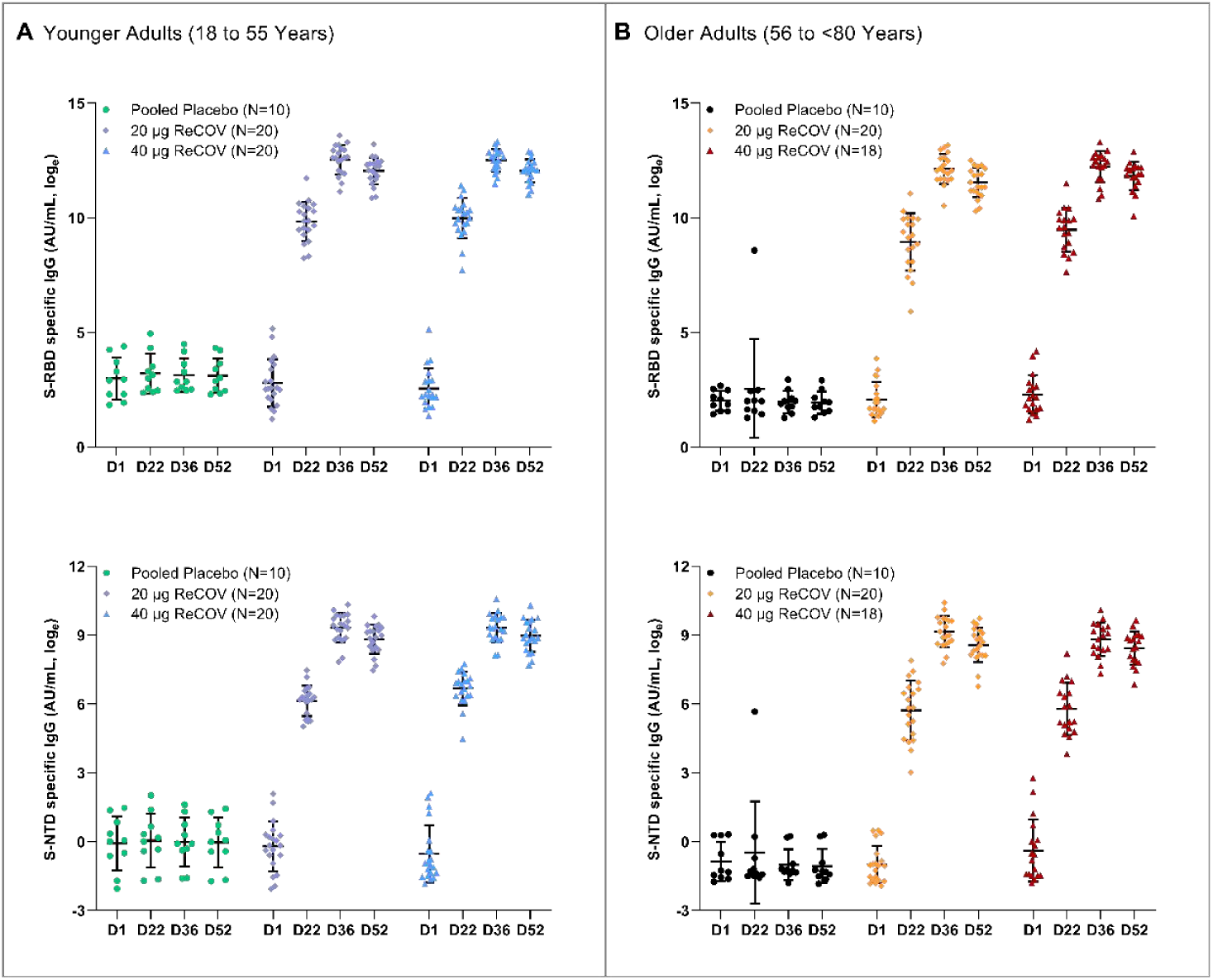
SARS-CoV-2 S-RBD and S-NTD specific IgG binding antibodies responses (GMT) after the first and second doses by age group. The specific IgG binding antibodies titers (GMT) of SARS-CoV-2 S-RBD and S-NTD against hCoV-19 Australia. The titers for S-RBD and S-NTD at Day 1 (D1), Day 22 (D22), Day 36 (D36) and Day 52 (D52) of vaccinated groups (pooled placebo, 20µg ReCOV, and 40µg ReCOV) were reported correspondingly for younger adults (Panel A) and older adults (Panel B) by natural base of log transformation.

Th1 biased cellular immune responses were observed after receiving ReCOV vaccination, irrespective of dose levels and subject ages (figure 5). In younger adults receiving 20 μg and 40 μg ReCOV, the average percentage of CD4^+^ T cells with IL-2 secretion was 0.01% and 0.02% at baseline, then reached 0.20% and 0.18% at Day 36, and remained 0.20% and 0.20% at Day 52, respectively. In addition, in younger adults receiving 20 μg and 40 μg ReCOV, the average percentage of CD4^+^ T cells with IFN-γ secretion was 0.01% and 0.01% at baseline, increased to 0.11% and 0.07% at Day 36, and remained at the same level at Day 52, respectively. Similar trends of CD4^+^ T with IL-2 and IFN-γ secretions were observed in older adults receiving 20 μg or 40 μg ReCOV vaccination. In contrast to increased secretion of Th1 cytokines, no obvious increased secretions of Th2 cytokines (IL-4 and IL-5) were observed in both age groups receiving 20 μg and 40 μg ReCOV.

**Figure 5:**
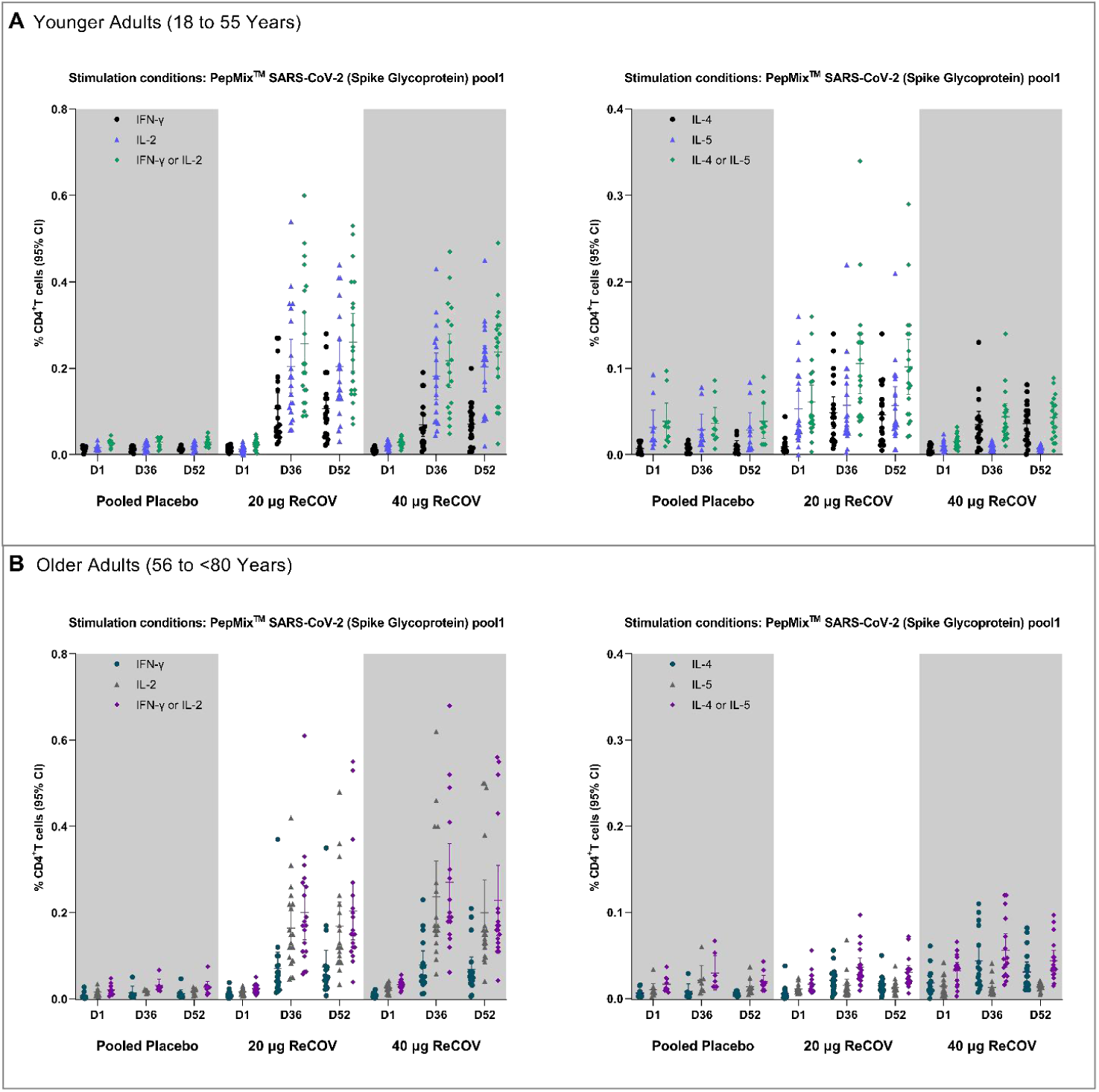
Th1 and Th2 cytokines responses after the first and second doses by age group. The percentage of CD4^+^ T cells for Th1 (IL-2, IFN-γ, IL-2 or IFN-γ) and Th2 (IL-4, IL-5, IL-4 or IL-5) cell responses at Day 1 (D1), Day 36 (D36) and Day 52 (D52) of vaccinated groups (pooled placebo, 20µg ReCOV, and 40µg ReCOV) were reported in younger adults (Panel A) and older adults (Panel B). Stimulation conditions: PepMix™ SARS-CoV-2 (Spike Glycoprotein) pool1.

## Discussion

Our data showed that ReCOV, a BFA03-adjuvanted recombinant 2-component subunit vaccine for COVID-19, at dose level of 20 μg and 40 μg, were well tolerated in healthy subjects aged 18-80 years, when administered as 2 intramuscular injections with 21 days interval. The safety profiles were similar between younger and older adults with both 20μg and 40μg ReCOV vaccination. Most solicited AEs were transient and mild in severity except for one participant who experienced moderate fever. The frequency of solicited AEs were generally higher after the second vaccination, which was in line with that reported for other protein subunit vaccines and mRNA vaccines.^11,12^

ReCOV is adjuvanted by BFA03, which is an AS03-like squalene adjuvant. The current phase I study indicated that ReCOV seemed to have a better safety profile comparing to other AS03 adjuvanted recombinant protein vaccines against COVID-19. In a phase I dose-ranging study of CoV2 preS dTM vaccine,^11^ a AS03 adjuvanted COVID-19 recombinant protein vaccine developed by Sanofi, 24% (20/85) and 13% (11/85) participants experienced grade 3 erythema and swelling in the high dose group. Additionally, 11% (9/80), 17% (13/79), and 14% (11/80) participants experienced grade 3 headache, malaise, and myalgia, respectively, in the low dose group. In a phase I trial of CoVLP, a plant-produced virus-like particle SARS-CoV-2 vaccine developed by Medicago, eight grade 3 solicited AEs were reported in five subjects received AS03-adjuvanted formulations after the second dose (8.5%, 5/59).^13^ In this study, the frequency of solicited AEs were generally higher after the second dosing. The same tendency has been reported for other protein subunit vaccines and mRNA vaccines. In the phase I study of CoV2 preS dTM, solicited injection-site and systemic reactions, including Grade 3 reactions, occurred more frequently after the second dosing than after the first dosing.^11^ In the phase I study of an AS03 adjuvanted recombinant protein vaccine candidate, SCB-2019, the frequency of systemic adverse events was 25-38% per group after the first dose and 44–56% after the second dose, with a concomitant increase in the proportion of Grade 2.^14^ Although the solicited adverse events observed in the current study were mild, close safety monitoring is warranted in future studies after the repeated ReCOV vaccination.

Good immunogenicity of ReCOV at both 20 μg and 40 μg were also well demonstrated in the younger and older adults. Strong SARS-CoV-2 neutralizing antibodies could be elicited by 20μg or 40μg ReCOV in most vaccinated subjects even after the first vaccination, and then peaked at 14 days and remained at high levels at 30 days after the second vaccination. The strong humoral immune responses were also demonstrated by 100% of SCR and high levels of GMTs for SARS-CoV-2 S-RBD and S-NTD specific IgG in subjects received ReCOV.

Humoral responses, especially neutralizing antibodies, have been considered as immune correlates of protection against SARS-CoV-2.^5^ Considering the diversity in laboratory testing methodologies and lack of head-to-head clinical trials, the neutralizing antibodies measured in this study were calibrated by WHO international standard which allows the conversion of the assay results to International Units (IU/mL). This enabled comparison of the results in the current study with immunogenicity data by other COVID-19 vaccines in a certain level. It is excited to observe that ReCOV elicited strong neutralizing antibodies with at least similar level to several COVID-19 vaccines with proved promising efficacies. As shown in both younger and older adults, 20 or 40μg ReCOV could elicit high level of neutralizing antibodies, with GMT of 1643.17∼1122.32 IU/mL, respectively. For two recombinant protein vaccines, SCB-2019 and MVC-COV1901, the GMTs of neutralizing antibodies post the second dosing were 224 IU/mL and 408 IU/mL, respectively. In addition, GMTs of neutralizing antibodies were 1404.16 IU/mL and 928.75 IU/mL from recipients of Moderna and Pfizer vaccines after two doses of vaccination.^15^ Therefore, the current study suggested that ReCOV may have high effective potential to protect occurrence of COVID-19 and prevent severe diseases. In addition, post hoc analysis indicated cross-neutralizing activities against Delta variants by both 20μg and 40μg ReCOV, with 5-11 times of reduction compared to the neutralization activity against wild-type SARS-CoV-2 (table S8), which showed similar reduction fold comparing with other COVID-19 vaccines.^14^

In addition to humoral response, Th1 cytokines were shown as the predominant phenotype after receiving ReCOV vaccination. A strong bias toward T-helper cell type 1 (Th1) phenotype was observed in both 20μg and 40μg ReCOV and in younger and older adult groups, indicated by INF-γ and IL-2 secretion in CD4^+^ T cell after antigen stimulation, while no obvious secretion of Th2 cytokines (e.g., IL-4 and IL-5) was observed. Th1 cytokines are important for development of T cell responses, CD4^+^ T cell is required for good induction of memory B cells. The similar trend of Th1 biased cellular immune responses have also been reported in studies on mRNA-1273^14^ and BNT162b2^16^ vaccine. Such a Th1 biased immune response is desirable for the development of a SARS-CoV-2 vaccine, due to the hypothetical concern for immune-mediated disease enhancement observed in preclinical studies for other coronaviruses.^17^

The current study showed that 40μg ReCOV, compared to the 20μg dose level, tended to induce more seroconversions and slightly higher level of neutralizing antibody after the first dose vaccination. However, 20μg ReCOV induced higher level of neutralizing antibodies than 40μg ReCOV after the second dosing. In addition, 20 μg or 40 μg ReCOV could elicit similar trends of CD4^+^ T with IL-2 and IFN-γ secretion, in both age groups. The appropriate dosage of ReCOV for primary and booster vaccination indications needs to be evaluated in further studies.

This study has few limitations. First, the study design did not contain an unadjuvanted group to measure the impact of BFA03 on the safety and immunogenicity, although the pre-clinical studies detected very limited immune response by ReCOV antigen alone, with around 300-fold lower compared to ReCOV.^7^ Second, the small sample size of the phase I study might not capture the rare occurred adverse events, the safety profile will need further evaluation in larger studies. Third, the primary analysis was performed till 30 days after the second vaccination, long-term safety and immune persistence need to be further evaluated after data collection of ongoing study follow-up. Last, the study mainly evaluated neutralizing antibodies against wild-type SARS-COV-2, although post-hoc analysis indicated cross-neutralizing activities against the Delta variant. Neutralizing activities against Omicron, was not tested due to unavailability of the assay in the central laboratory yet. Limited neutralizing activity against Omicron induced by primary immunization with all other COVID-19 vaccines in market designed for the wild-type strain, as shown that after the primary two-dose series of the mRNA-1273 vaccine, neutralization titers against Omicron variant were 35.0 times lower than those against D614G variant, indicating an increased risk of severe breakthrough infection,^18^ also there was a significant reduction in GMT of hACE2 receptor binding inhibition against Omicron variant compared to the ancestral strain after primary vaccination by NVX-CoV2373.^19^ Although the potential cross-neutralization against Omicron could be expected for ReCOV, based on its RBD-NTD-foldon design with more conservative antigen epitopes, the decreasing of neutralizing antibodies might result in an ineffective protection against Omicron. It suggested that clinical trials for ReCOV with booster vaccination indications, including both homologous and heterologous boosting should be speeded up as well, to achieve more evidences for the appropriate immunization schedules for ReCOV.

In conclusion, this phase I study demonstrated good safety profile and strong immunogenicity of ReCOV in the study population aged between 18-55 years or 56-80 years. The available data strongly supports the accelerated development of ReCOV.

## Panel: Research in context

### Evidence before this study

In response to the worldwide pandemic induced by the severe acute respiratory syndrome coronavirus 2 (SARS-CoV-2), till now there are 10 vaccines have been listed by world health organization (WHO) for emergency use, with 2 subunit protein vaccines included. ReCOV, the first recombinant trimeric NTD and RBD two-component SARS-CoV-2 subunit vaccine adjuvanted with BFA03, has previously been reported to be immunogenic and protective against pneumonia in hACE2 transgenic mice and rhesus macaque challenge models. We searched in PubMed for research articles published between database inception and March 31, 2022, using the terms ‘COVID-19’, ‘SARS-CoV-2’, ‘vaccine’, ‘clinical trial’, and ‘subunit’. No language restrictions were applied. We identified six studies on human clinical trials of SARS-CoV-2 subunit vaccines, including SCB-2019, CoVLP, AKS-452, CoVac-1 and Sclamp. All these subunit vaccines clinical trials revealed good tolerability and safety of studied vaccines, four of which reported data of both humoral and cellular immunogenicity, while the ability to induce neutralizing antibodies against the SARS-CoV-2 variants varied from each other. Two studies compared different adjuvants, SCB-2019 formulated with AS03 and CpG/Alum adjuvants, and CoVLP with AS03 and CpG1018 adjuvants. Both SCB-2019 and CoVLP induced higher levels of antibodies and cellular immune responses when adjuvanted with AS03.

### Added value of this study

We reported the results of the first clinical study of ReCOV in both young and older subjects. The ReCOV vaccine was safe and tolerated after two-dose vaccination in both younger and older adults. Good immunogenicity of ReCOV was well demonstrated, the strong SARS-CoV-2 neutralizing antibodies could be elicited both 20 μg and 40 μg ReCOV in majority vaccinated subjects even after the first vaccination, then further peaked at 14 days post the second vaccination. The strong humoral immune responses were also demonstrated by 100% seroconversion rate and high levels of SARS-CoV-2 S-RBD and S-NTD specific IgG in subjects received ReCOV. Furthermore, Th1 biased cellular immune responses were observed after receiving ReCOV vaccination, irrespective of dose levels and subject ages.

### Implications of all the available evidence

The available safety and immunogenicity data support the progression of following trials for ReCOV. Besides of the well safety profile, immunization with ReCOV results in rapid induction of both humoral and cellular immune responses against SARS-CoV-2, with increased responses after a second dose. Further clinical studies, including efficacy assessment and for boosting, should be done with this investigational vaccine, relevant studies, e.g., NCT05084989 study with two-dose primary vaccination, and NCT05323435 study with one dose booster vaccination are ongoing.

## Data Availability

All data produced in the present study are available upon reasonable request to the authors

## Contributors

Chris Wynne and Paul Hamilton are the principal investigators of this trial. Jing-Xin Li and Feng-Cai Zhu contributed to clinical development plan and study protocol key design. Zijing Yue and Chen Mo drafted of the manuscript. Jing-Xin Li, Jian-Hui Zhang and Kun-Xue Hong contributed to critical review and revising of the manuscript. Jianhui Zhang, Jianping Chen, Yong Liu, Jing-Xin Li and Fengcai Zhu contributed to study supervision. Jia-Ping Yu and Wen-Rong Yao contributed to the design of the investigational vaccine. All authors reviewed and approved the final manuscript.

## Data sharing

Data Sharing Statement will be available with the full text of this article upon publication. https://clinicaltrials.gov/ct2/show/NCT04818801

## Acknowledgment

We are grateful for all investigators at the Auckland Clinical Studies and Christchurch Clinical Studies Trust, New Zealand who contributed to the site work of the trial; We are grateful for all investigators at the central laboratory 360Biolabs, Australia who performed laboratory tests of the trial. We thank all the participants in the trial for their dedication to the trial.

