## Supplemental Table 1 for "The preliminary safety and immunogenicity results of a randomized, double-blind, placebo-controlled Phase I trial for a recombinant two-component subunit SARS-CoV-2 vaccine ReCOV"

Table S1 Overview of Adverse Events up to 30 Days After the Second Dose (Safety Analysis Set)

| **Status** | **18 to 55 years** | | | | **56 to <80 years** | | |
| --- | --- | --- | --- | --- | --- | --- | --- |
|  | **Pooled Placebo (N=10) n (%) E** | **20μg ReCOV (N=20) n (%) E** | **40μg ReCOV (N=20) n (%) E** | **Pooled Placebo (N=10) n (%) E** | | **20μg ReCOV (N=20) n (%) E** | **40μg ReCOV (N=19) n (%) E** |
| **At least one TEAE** | 4 (40.0) 6 | 18 (90.0) 90 | 18 (90.0) 82 | 8 (80.0) 30 | | 16 (80.0) 64 | 18 (94.7) 84 |
| **At least one related TEAE** | 3 (30.0) 3 | 16 (80.0) 75 | 18 (90.0) 61 | 5 (50.0) 15 | | 13 (65.0) 51 | 17 (89.5) 74 |
| **At least one solicited TEAE** | 4 (40.0) 4 | 14 (70.0) 67 | 16 (80.0) 48 | 5 (50.0) 10 | | 13 (65.0) 42 | 16 (84.2) 63 |
| **At least one solicited local TEAE** | 1 (10.0) 1 | 12 (60.0) 30 | 14 (70.0) 25 | 1 (10.0) 2 | | 11 (55.0) 19 | 16 (84.2) 27 |
| **At least one solicited systemic TEAE** | 3 (30.0) 3 | 12 (60.0) 37 | 12 (60.0) 23 | 5 (50.0) 8 | | 10 (50.0) 23 | 10 (52.6) 36 |
| **At least one related solicited systemic TEAE** | 2 (20.0) 2 | 12 (60.0) 36 | 12 (60.0) 23 | 5 (50.0) 7 | | 10 (50.0) 23 | 10 (52.6) 35 |
| **At least one injection site reaction** | 1 (10.0) 1 | 12 (60.0) 31 | 14 (70.0) 26 | 2 (20.0) 3 | | 12 (60.0) 20 | 17 (89.5) 30 |
| **At least one unsolicited TEAE** | 2 (20.0) 2 | 14 (70.0) 23 | 12 (60.0) 34 | 7 (70.0) 20 | | 11 (55.0) 22 | 12 (63.2) 21 |
| **At least one related unsolicited TEAE** | 0 | 8 (40.0) 9 | 9 (45.0) 13 | 2 (20.0) 6 | | 7 (35.0) 9 | 8 (42.1) 12 |
| **At least one Grade ≥3 unsolicited TEAE** | 0 | 0 | 0 | 0 | | 0 | 0 |
| **At least one Grade ≥3 related unsolicited TEAE** | 0 | 0 | 0 | 0 | | 0 | 0 |
| **At least one serious TEAE** | 0 | 0 | 0 | 0 | | 0 | 0 |
| **At least one related serious TEAE** | 0 | 0 | 0 | 0 | | 0 | 0 |
| **At least one TEAE of special interest** | 0 | 0 | 0 | 0 | | 0 | 0 |
| **At least one TEAE meeting the stopping criteria** | 0 | 0 | 0 | 0 | | 0 | 0 |
| **At least one TEAE leading to death** | 0 | 0 | 0 | 0 | | 0 | 0 |
| **At least one TEAE leading to study discontinuation** | 0 | 0 | 0 | 0 | | 0 | 0 |
| **At least one TEAE leading to IP discontinuation** | 0 | 0 | 0 | 0 | | 0 | 0 |
| **TEAE by Severity:** |  |  |  |  | |  |  |
| Mild (Grade 1) | 4 (40.0) 6 | 18 (90.0) 88 | 17 (85.0) 80 | 8 (80.0) 27 | | 16 (80.0) 64 | 18 (94.7) 84 |
| Moderate (Grade 2) | 0 | 2 (10.0) 2 | 2 (10.0) 2 | 2 (20.0) 3 | | 0 | 0 |
| Severe (Grade 3) | 0 | 0 | 0 | 0 | | 0 | 0 |
| Potentially life-threatening (Grade 4) | 0 | 0 | 0 | 0 | | 0 | 0 |
| **IP-related TEAE by Severity:** |  |  |  |  | |  |  |
| Mild (Grade 1) | 3 (30.0) 3 | 16 (80.0) 74 | 17 (85.0) 59 | 5 (50.0) 15 | | 13 (65.0) 51 | 17 (89.5) 74 |
| Moderate (Grade 2) | 0 | 1 (5.0) 1 | 2 (10.0) 2 | 0 | | 0 | 0 |
| Severe (Grade 3) | 0 | 0 | 0 | 0 | | 0 | 0 |
| Potentially life-threatening (Grade 4) | 0 | 0 | 0 | 0 | | 0 | 0 |

AE: Adverse Event; IP: Investigational Product; MedDRA: Medical Dictionary for Regulatory Activities; TEAE: Treatment-Emergent AE.

N = Total number of subjects in the relevant analysis set. n = Number of subjects with at least one TEAE in each category (subjects with multiple AEs in each category are counted only once in each category). % = Percentage of subjects in each category calculated relative to the total number of subjects in the relevant analysis set. E = Number of events in each category.

TEAEs are defined as AEs that started or worsened in severity on or after the date of first dose of IP up to 30 days after the second dose.

Solicited local TEAEs are defined as AEs coded to ‘Injection site pain’, ‘Injection site erythema’ (redness) or ’Injection site swelling’ up to 7 days after each dose. Solicited systemic TEAEs are defined as AEs coded to ‘Fatigue’, ‘Pyrexia’ (fever), ‘Nausea’, ‘Vomiting’, ‘Diarrhea’, ‘Headache’ or ‘Myalgia’(muscle pain) up to 7 days after each dose.

Injection site reactions are defined as related AEs coded to the High Level Group Term 'Administration site reactions' up to 7 days after each dose.

Unsolicited TEAEs are defined as AEs that started up to 30 days after the second dose, excluding solicited local and systemic TEAEs up to 7 days after each dose. Related AEs are defined as AEs with a relationship to IP classed as 'Possible', 'Probable' or 'Certain' and includes events with a missing relationship. Serious AEs are defined as any AE for which 'Serious event' was indicated as 'Yes'.

AEs of Special Interest are defined as AEs that include potential immune mediated diseases, as well as COVID-19 vaccine and COVID-19 illness related AEs. Stopping criteria AEs are defined as any AE for which ‘Did this event meet stopping criteria as per protocol’ was indicated as ‘Yes’.

Date source: Table 14.3.1.1.1.
