## Supplemental Table 2 for "The preliminary safety and immunogenicity results of a randomized, double-blind, placebo-controlled Phase I trial for a recombinant two-component subunit SARS-CoV-2 vaccine ReCOV"

Table S2 Frequency of Solicited Local Adverse Events up to 7 Days Following Each Vaccination (Safety Analysis Set)

| **Preferred**  **Term** | **Most Recent Dose** | **18 to 55 years** | | | | **56 to <80 years** | | |
| --- | --- | --- | --- | --- | --- | --- | --- | --- |
|  |  | **Pooled Placebo (N=10) n (%) E** | **20μg ReCOV (N=20) n (%) E** | **40μg ReCOV (N=20) n (%) E** | **Pooled Placebo (N=10) n (%) E** | | **20μg ReCOV (N=20) n (%) E** | **40μg ReCOV (N=19) n (%) E** |
| **At least one solicited local TEAE** | First Dose | 1 (10.0) 1 | 10 (50.0) 12 | 10 (50.0) 10 | 1 (10.0) 2 | | 8 (40.0) 8 | 10 (52.6) 11 |
|  | Second Dose | 0 | 11 (55.0) 18 | 13 (65.0) 15 | 0 | | 10 (50.0) 11 | 12 (63.2) 16 |
| **Injection site pain** | First Dose | 1 (10.0) 1 | 9 (45.0) 9 | 10 (50.0) 10 | 1 (10.0) 1 | | 6 (30.0) 6 | 9 (47.4) 10 |
|  | Second Dose | 0 | 10 (50.0) 10 | 13 (65.0) 13 | 0 | | 9 (45.0) 9 | 11 (57.9) 12 |
| **Injection site swelling** | First Dose | 0 | 3 (15.0) 3 | 0 | 1 (10.0) 1 | | 2 (10.0) 2 | 1 (5.3) 1 |
|  | Second Dose | 0 | 6 (30.0) 6 | 2 (10.0) 2 | 0 | | 2 (10.0) 2 | 3 (15.8) 3 |
| **Injection site erythema** | First Dose | 0 | 0 | 0 | 0 | | 0 | 0 |
|  | Second Dose | 0 | 2 (10.0) 2 | 0 | 0 | | 0 | 1 (5.3) 1 |

AE: Adverse Event; MedDRA: Medical Dictionary for Regulatory Activities; TEAE: Treatment-Emergent AE.

N = Total number of subjects in the relevant analysis set.

n = Number of subjects with at least one TEAE in each category (subjects with multiple AEs in each category are counted only once in each category).

% = Percentage of subjects in each category calculated relative to the total number of subjects in the relevant analysis set. E = Number of events in each category.

Solicited local TEAEs are defined as AEs coded to ‘Injection site pain’, ‘Injection site erythema’ (redness) or ’Injection site swelling’ up to 7 days after each dose. AEs were coded using MedDRA Version 24.0.

Date source: Table 14.3.1.2.2.
