## Supplemental Table 3 for "The preliminary safety and immunogenicity results of a randomized, double-blind, placebo-controlled Phase I trial for a recombinant two-component subunit SARS-CoV-2 vaccine ReCOV"

| Preferred Term (unit)  Most Recent Dose | Statistic | Pooled Placebo (N=10) n (%) | 20 ug ReCOV (N=20) n (%) | 40 ug ReCOV (N=20) n (%) | Pooled ReCOV (N=40) n (%) | Overall (N=50) n (%) |
| --- | --- | --- | --- | --- | --- | --- |
| Injection site pain (days) | | | | | | |
| First Dose | n | 1 | 9 | 10 | 19 | 20 |
|  | Mean | 2.0 | 3.7 | 2.7 | 3.2 | 3.1 |
|  | SD | - | 2.12 | 0.95 | 1.64 | 1.62 |
|  | Median | 2.0 | 3.0 | 3.0 | 3.0 | 3.0 |
|  | Minimum | 2 | 1 | 1 | 1 | 1 |
|  | Maximum | 2 | 8 | 4 | 8 | 8 |
| Second Dose | n | 0 | 10 | 13 | 23 | 23 |
|  | Mean | - | 3.8 | 3.8 | 3.8 | 3.8 |
|  | SD | - | 1.55 | 1.24 | 1.35 | 1.35 |
|  | Median | - | 4.0 | 4.0 | 4.0 | 4.0 |
|  | Minimum | - | 2 | 2 | 2 | 2 |
|  | Maximum | - | 7 | 6 | 7 | 7 |

| Preferred Term (unit)  Most Recent Dose | Statistic | Pooled Placebo (N=10) n (%) | 20 ug ReCOV (N=20) n (%) | 40 ug ReCOV (N=20) n (%) | Pooled ReCOV (N=40) n (%) | Overall (N=50) n (%) |
| --- | --- | --- | --- | --- | --- | --- |
| Injection site swelling (days) | | | | | | |
| First Dose | n | 0 | 3 | 0 | 3 | 3 |
|  | Mean | - | 2.3 | - | 2.3 | 2.3 |
|  | SD | - | 1.15 | - | 1.15 | 1.15 |
|  | Median | - | 3.0 | - | 3.0 | 3.0 |
|  | Minimum | - | 1 | - | 1 | 1 |
|  | Maximum | - | 3 | - | 3 | 3 |
| Second Dose | n | 0 | 6 | 2 | 8 | 8 |
|  | Mean | - | 3.2 | 3.0 | 3.1 | 3.1 |
|  | SD | - | 0.75 | 1.41 | 0.83 | 0.83 |
|  | Median | - | 3.0 | 3.0 | 3.0 | 3.0 |
|  | Minimum | - | 2 | 2 | 2 | 2 |
|  | Maximum | - | 4 | 4 | 4 | 4 |
| Injection site erythema (days) | | | | | | |
| Second Dose | n | 0 | 2 | 0 | 2 | 2 |
|  | Mean | - | 3.5 | - | 3.5 | 3.5 |
|  | SD | - | 0.71 | - | 0.71 | 0.71 |
|  | Median | - | 3.5 | - | 3.5 | 3.5 |
|  | Minimum | - | 3 | - | 3 | 3 |
|  | Maximum | - | 4 | - | 4 | 4 |

| Preferred Term (unit)  Most Recent Dose | Statistic | Pooled Placebo (N=10) n (%) | 20 ug ReCOV (N=20) n (%) | 40 ug ReCOV (N=19) n (%) | Pooled ReCOV (N=39) n (%) | Overall (N=49) n (%) |
| --- | --- | --- | --- | --- | --- | --- |
| Injection site pain (days) | | | | | | |
| First Dose | n | 1 | 6 | 9 | 15 | 16 |
|  | Mean | 2.0 | 2.8 | 10.3 | 7.3 | 7.0 |
|  | SD | - | 1.83 | 14.38 | 11.57 | 11.25 |
|  | Median | 2.0 | 3.0 | 3.0 | 3.0 | 3.0 |
|  | Minimum | 2 | 1 | 2 | 1 | 1 |
|  | Maximum | 2 | 6 | 38 | 38 | 38 |
| Second Dose | n | 0 | 9 | 11 | 20 | 20 |
|  | Mean | - | 5.3 | 3.7 | 4.5 | 4.5 |
|  | SD | - | 6.44 | 2.72 | 4.70 | 4.70 |
|  | Median | - | 3.0 | 3.0 | 3.0 | 3.0 |
|  | Minimum | - | 2 | 1 | 1 | 1 |
|  | Maximum | - | 22 | 11 | 22 | 22 |

| Preferred Term (unit)  Most Recent Dose | Statistic | Pooled Placebo (N=10) n (%) | 20 ug ReCOV (N=20) n (%) | 40 ug ReCOV (N=19) n (%) | Pooled ReCOV (N=39) n (%) | Overall (N=49) n (%) |
| --- | --- | --- | --- | --- | --- | --- |
| Injection site swelling (days) | | | | | | |
| First Dose | n | 1 | 2 | 1 | 3 | 4 |
|  | Mean | 2.0 | 12.0 | 3.0 | 9.0 | 7.3 |
|  | SD | - | 14.14 | - | 11.27 | 9.84 |
|  | Median | 2.0 | 12.0 | 3.0 | 3.0 | 2.5 |
|  | Minimum | 2 | 2 | 3 | 2 | 2 |
|  | Maximum | 2 | 22 | 3 | 22 | 22 |
| Second Dose | n | 0 | 2 | 3 | 5 | 5 |
|  | Mean | - | 3.5 | 2.0 | 2.6 | 2.6 |
|  | SD | - | 2.12 | 1.00 | 1.52 | 1.52 |
|  | Median | - | 3.5 | 2.0 | 2.0 | 2.0 |
|  | Minimum | - | 2 | 1 | 1 | 1 |
|  | Maximum | - | 5 | 3 | 5 | 5 |
| Injection site erythema (days) | | | | | | |
| Second Dose | n | 0 | 0 | 1 | 1 | 1 |
|  | Mean | - | - | 3.0 | 3.0 | 3.0 |
|  | SD | - | - | - | - | - |
|  | Median | - | - | 3.0 | 3.0 | 3.0 |
|  | Minimum | - | - | 3 | 3 | 3 |
|  | Maximum | - | - | 3 | 3 | 3 |

| Preferred Term (unit)  Most Recent Dose | Statistic | Pooled Placebo (N=20) n (%) | 20 ug ReCOV (N=40) n (%) | 40 ug ReCOV (N=39) n (%) | Pooled ReCOV (N=79) n (%) | Overall (N=99) n (%) |
| --- | --- | --- | --- | --- | --- | --- |
| Injection site pain (days) | | | | | | |
| First Dose | n | 2 | 15 | 19 | 34 | 36 |
|  | Mean | 2.0 | 3.3 | 6.3 | 5.0 | 4.8 |
|  | SD | 0.00 | 1.99 | 10.38 | 7.92 | 7.72 |
|  | Median | 2.0 | 3.0 | 3.0 | 3.0 | 3.0 |
|  | Minimum | 2 | 1 | 1 | 1 | 1 |
|  | Maximum | 2 | 8 | 38 | 38 | 38 |
| Second Dose | n | 0 | 19 | 24 | 43 | 43 |
|  | Mean | - | 4.5 | 3.8 | 4.1 | 4.1 |
|  | SD | - | 4.50 | 2.01 | 3.32 | 3.32 |
|  | Median | - | 4.0 | 3.5 | 4.0 | 4.0 |
|  | Minimum | - | 2 | 1 | 1 | 1 |
|  | Maximum | - | 22 | 11 | 22 | 22 |

| Preferred Term (unit)  Most Recent Dose | Statistic | Pooled Placebo (N=20) n (%) | 20 ug ReCOV (N=40) n (%) | 40 ug ReCOV (N=39) n (%) | Pooled ReCOV (N=79) n (%) | Overall (N=99) n (%) |
| --- | --- | --- | --- | --- | --- | --- |
| Injection site swelling (days) | | | | | | |
| First Dose | n | 1 | 5 | 1 | 6 | 7 |
|  | Mean | 2.0 | 6.2 | 3.0 | 5.7 | 5.1 |
|  | SD | - | 8.87 | - | 8.04 | 7.47 |
|  | Median | 2.0 | 3.0 | 3.0 | 3.0 | 3.0 |
|  | Minimum | 2 | 1 | 3 | 1 | 1 |
|  | Maximum | 2 | 22 | 3 | 22 | 22 |
| Second Dose | n | 0 | 8 | 5 | 13 | 13 |
|  | Mean | - | 3.3 | 2.4 | 2.9 | 2.9 |
|  | SD | - | 1.04 | 1.14 | 1.12 | 1.12 |
|  | Median | - | 3.0 | 2.0 | 3.0 | 3.0 |
|  | Minimum | - | 2 | 1 | 1 | 1 |
|  | Maximum | - | 5 | 4 | 5 | 5 |
| Injection site erythema (days) | | | | | | |
| Second Dose | n | 0 | 2 | 1 | 3 | 3 |
|  | Mean | - | 3.5 | 3.0 | 3.3 | 3.3 |
|  | SD | - | 0.71 | - | 0.58 | 0.58 |
|  | Median | - | 3.5 | 3.0 | 3.0 | 3.0 |
|  | Minimum | - | 3 | 3 | 3 | 3 |
|  | Maximum | - | 4 | 3 | 4 | 4 |
