## Supplemental Table 4 for "The preliminary safety and immunogenicity results of a randomized, double-blind, placebo-controlled Phase I trial for a recombinant two-component subunit SARS-CoV-2 vaccine ReCOV"

Table S4 Frequency of Solicited Systemic Treatment-Emergent Adverse Events up to 7 Days Following Each Vaccination (Safety Analysis Set)

| **Preferred Term** | **Most Recent Dose** | **18 to 55 years** | | | **56 to <80 years** | | | |
| --- | --- | --- | --- | --- | --- | --- | --- | --- |
|  |  | **Pooled Placebo (N=10) n (%) E** | **20μg ReCOV (N=20) n (%) E** | **40μg ReCOV (N=20) n (%) E** | | **Pooled Placebo (N=10) n (%) E** | **20μg ReCOV (N=20) n (%) E** | **40μg ReCOV (N=19) n (%) E** |
| **At least one solicited systemic TEAE** | First Dose | 1 (10.0) 1 | 5 (25.0) 5 | 4 (20.0) 5 | | 4 (40.0) 4 | 2 (10.0) 2 | 6 (31.6) 13 |
|  | Second Dose | 2 (20.0) 2 | 12 (60.0) 32 | 10 (50.0) 18 | | 3 (30.0) 4 | 10 (50.0) 21 | 8 (42.1) 23 |
| **Fatigue** | First Dose | 1 (10.0) 1 | 3 (15.0) 3 | 2 (10.0) 2 | | 1 (10.0) 1 | 2 (10.0) 2 | 4 (21.1) 4 |
|  | Second Dose | 0 | 10 (50.0) 10 | 3 (15.0) 3 | | 2 (20.0) 2 | 7 (35.0) 7 | 7 (36.8) 7 |
| **Headache** | First Dose | 0 | 1 (5.0) 1 | 2 (10.0) 2 | | 1 (10.0) 1 | 0 | 4 (21.1) 4 |
|  | Second Dose | 1 (10.0) 1 | 8 (40.0) 8 | 5 (25.0) 6 | | 2 (20.0) 2 | 4 (20.0) 4 | 5 (26.3) 6 |
| **Myalgia** | First Dose | 0 | 1 (5.0) 1 | 1 (5.0) 1 | | 1 (10.0) 1 | 0 | 3 (15.8) 3 |
|  | Second Dose | 0 | 7 (35.0) 7 | 6 (30.0) 6 | | 0 | 7 (35.0) 8 | 4 (21.1) 4 |
| **Nausea** | First Dose | 0 | 0 | 0 | | 1 (10.0) 1 | 0 | 1 (5.3) 1 |
|  | Second Dose | 0 | 2 (10.0) 2 | 3 (15.0) 3 | | 0 | 0 | 2 (10.5) 2 |
| **Pyrexia** | First Dose | 0 | 0 | 0 | | 0 | 0 | 0 |
|  | Second Dose | 0 | 4 (20.0) 4 | 0 | | 0 | 2 (10.0) 2 | 4 (21.1) 4 |
| **Diarrhoea** | First Dose | 0 | 0 | 0 | | 0 | 0 | 1 (5.3) 1 |
|  | Second Dose | 1 (10.0) 1 | 0 | 0 | | 0 | 0 | 0 |
| **Vomiting** | First Dose | 0 | 0 | 0 | | 0 | 0 | 0 |
|  | Second Dose | 0 | 1 (5.0) 1 | 0 | | 0 | 0 | 0 |

AE: Adverse Event; MedDRA: Medical Dictionary for Regulatory Activities; TEAE: Treatment-Emergent AE.

N = Total number of subjects in the relevant analysis set.

n = Number of subjects with at least one TEAE in each category (subjects with multiple AEs in each category are counted only once in each category).

% = Percentage of subjects in each category calculated relative to the total number of subjects in the relevant analysis set. E = Number of events in each category.

Solicited systemic TEAEs are defined as AEs coded to ‘Fatigue’, ‘Pyrexia’ (fever), ‘Nausea’, ‘Vomiting’, ‘Diarrhoea‘, ‘Headache’ or ‘Myalgia’(muscle pain) up to 7 days after each dose. AEs were coded using MedDRA Version 24.0.

Date source: Table 14.3.1.2.3.
