## Supplemental Table 5 for "The preliminary safety and immunogenicity results of a randomized, double-blind, placebo-controlled Phase I trial for a recombinant two-component subunit SARS-CoV-2 vaccine ReCOV"

| Preferred Term (unit)  Most Recent Dose | Statistic | Pooled Placebo (N=10) n (%) | 20 ug ReCOV (N=20) n (%) | 40 ug ReCOV (N=20) n (%) | Pooled ReCOV (N=40) n (%) | Overall (N=50) n (%) |
| --- | --- | --- | --- | --- | --- | --- |
| Fatigue (days) | | | | | | |
| First Dose | n | 1 | 3 | 2 | 5 | 6 |
|  | Mean | 7.0 | 2.7 | 3.0 | 2.8 | 3.5 |
|  | SD | - | 1.15 | 1.41 | 1.10 | 1.97 |
|  | Median | 7.0 | 2.0 | 3.0 | 2.0 | 3.0 |
|  | Minimum | 7 | 2 | 2 | 2 | 2 |
|  | Maximum | 7 | 4 | 4 | 4 | 7 |
| Second Dose | n | 0 | 10 | 3 | 13 | 13 |
|  | Mean | - | 2.9 | 4.3 | 3.2 | 3.2 |
|  | SD | - | 2.60 | 3.21 | 2.68 | 2.68 |
|  | Median | - | 2.0 | 3.0 | 2.0 | 2.0 |
|  | Minimum | - | 1 | 2 | 1 | 1 |
|  | Maximum | - | 10 | 8 | 10 | 10 |

| Preferred Term (unit)  Most Recent Dose | Statistic | Pooled Placebo (N=10) n (%) | 20 ug ReCOV (N=20) n (%) | 40 ug ReCOV (N=20) n (%) | Pooled ReCOV (N=40) n (%) | Overall (N=50) n (%) |
| --- | --- | --- | --- | --- | --- | --- |
| Headache (days) | | | | | | |
| First Dose | n | 0 | 1 | 2 | 3 | 3 |
|  | Mean | - | 3.0 | 3.0 | 3.0 | 3.0 |
|  | SD | - | - | 2.83 | 2.00 | 2.00 |
|  | Median | - | 3.0 | 3.0 | 3.0 | 3.0 |
|  | Minimum | - | 3 | 1 | 1 | 1 |
|  | Maximum | - | 3 | 5 | 5 | 5 |
| Second Dose | n | 1 | 8 | 5 | 13 | 14 |
|  | Mean | 1.0 | 3.1 | 2.4 | 2.8 | 2.7 |
|  | SD | - | 2.85 | 0.55 | 2.23 | 2.20 |
|  | Median | 1.0 | 2.0 | 2.0 | 2.0 | 2.0 |
|  | Minimum | 1 | 1 | 2 | 1 | 1 |
|  | Maximum | 1 | 10 | 3 | 10 | 10 |

| Preferred Term (unit)  Most Recent Dose | Statistic | Pooled Placebo (N=10) n (%) | 20 ug ReCOV (N=20) n (%) | 40 ug ReCOV (N=20) n (%) | Pooled ReCOV (N=40) n (%) | Overall (N=50) n (%) |
| --- | --- | --- | --- | --- | --- | --- |
| Myalgia (days) | | | | | | |
| First Dose | n | 0 | 1 | 1 | 2 | 2 |
|  | Mean | - | 2.0 | 4.0 | 3.0 | 3.0 |
|  | SD | - | - | - | 1.41 | 1.41 |
|  | Median | - | 2.0 | 4.0 | 3.0 | 3.0 |
|  | Minimum | - | 2 | 4 | 2 | 2 |
|  | Maximum | - | 2 | 4 | 4 | 4 |
| Second Dose | n | 0 | 7 | 6 | 13 | 13 |
|  | Mean | - | 3.7 | 2.7 | 3.2 | 3.2 |
|  | SD | - | 2.63 | 0.82 | 2.01 | 2.01 |
|  | Median | - | 3.0 | 2.5 | 3.0 | 3.0 |
|  | Minimum | - | 1 | 2 | 1 | 1 |
|  | Maximum | - | 9 | 4 | 9 | 9 |
| Nausea (days) | | | | | | |
| Second Dose | n | 0 | 2 | 3 | 5 | 5 |
|  | Mean | - | 3.5 | 2.7 | 3.0 | 3.0 |
|  | SD | - | 3.54 | 1.15 | 2.00 | 2.00 |
|  | Median | - | 3.5 | 2.0 | 2.0 | 2.0 |
|  | Minimum | - | 1 | 2 | 1 | 1 |
|  | Maximum | - | 6 | 4 | 6 | 6 |

| Preferred Term (unit)  Most Recent Dose | Statistic | Pooled Placebo (N=10) n (%) | 20 ug ReCOV (N=20) n (%) | 40 ug ReCOV (N=20) n (%) | Pooled ReCOV (N=40) n (%) | Overall (N=50) n (%) |
| --- | --- | --- | --- | --- | --- | --- |
| Pyrexia (days) | | | | | | |
| Second Dose | n | 0 | 4 | 0 | 4 | 4 |
|  | Mean | - | 3.5 | - | 3.5 | 3.5 |
|  | SD | - | 2.52 | - | 2.52 | 2.52 |
|  | Median | - | 3.0 | - | 3.0 | 3.0 |
|  | Minimum | - | 1 | - | 1 | 1 |
|  | Maximum | - | 7 | - | 7 | 7 |
| Diarrhoea (days) | | | | | | |
| Second Dose | n | 1 | 0 | 0 | 0 | 1 |
|  | Mean | 2.0 | - | - | - | 2.0 |
|  | SD | - | - | - | - | - |
|  | Median | 2.0 | - | - | - | 2.0 |
|  | Minimum | 2 | - | - | - | 2 |
|  | Maximum | 2 | - | - | - | 2 |
| Vomiting (days) | | | | | | |
| Second Dose | n | 0 | 1 | 0 | 1 | 1 |
|  | Mean | - | 3.0 | - | 3.0 | 3.0 |
|  | SD | - | - | - | - | - |
|  | Median | - | 3.0 | - | 3.0 | 3.0 |
|  | Minimum | - | 3 | - | 3 | 3 |
|  | Maximum | - | 3 | - | 3 | 3 |

| Preferred Term (unit)  Most Recent Dose | Statistic | Pooled Placebo (N=10) n (%) | 20 ug ReCOV (N=20) n (%) | 40 ug ReCOV (N=19) n (%) | Pooled ReCOV (N=39) n (%) | Overall (N=49) n (%) |
| --- | --- | --- | --- | --- | --- | --- |
| Fatigue (days) | | | | | | |
| First Dose | n | 1 | 2 | 4 | 6 | 7 |
|  | Mean | 33.0 | 4.0 | 12.8 | 9.8 | 13.1 |
|  | SD | - | 2.83 | 13.52 | 11.48 | 13.66 |
|  | Median | 33.0 | 4.0 | 6.5 | 6.0 | 6.0 |
|  | Minimum | 33 | 2 | 5 | 2 | 2 |
|  | Maximum | 33 | 6 | 33 | 33 | 33 |
| Second Dose | n | 2 | 7 | 7 | 14 | 16 |
|  | Mean | 5.0 | 5.1 | 7.7 | 6.4 | 6.3 |
|  | SD | 1.41 | 6.23 | 11.35 | 8.90 | 8.31 |
|  | Median | 5.0 | 2.0 | 3.0 | 2.5 | 3.5 |
|  | Minimum | 4 | 2 | 2 | 2 | 2 |
|  | Maximum | 6 | 19 | 33 | 33 | 33 |

| Preferred Term (unit)  Most Recent Dose | Statistic | Pooled Placebo (N=10) n (%) | 20 ug ReCOV (N=20) n (%) | 40 ug ReCOV (N=19) n (%) | Pooled ReCOV (N=39) n (%) | Overall (N=49) n (%) |
| --- | --- | --- | --- | --- | --- | --- |
| Myalgia (days) | | | | | | |
| First Dose | n | 1 | 0 | 3 | 3 | 4 |
|  | Mean | 11.0 | - | 5.0 | 5.0 | 6.5 |
|  | SD | - | - | 1.73 | 1.73 | 3.32 |
|  | Median | 11.0 | - | 6.0 | 6.0 | 6.0 |
|  | Minimum | 11 | - | 3 | 3 | 3 |
|  | Maximum | 11 | - | 6 | 6 | 11 |
| Second Dose | n | 0 | 7 | 4 | 11 | 11 |
|  | Mean | - | 7.3 | 2.0 | 5.4 | 5.4 |
|  | SD | - | 11.40 | 0.82 | 9.23 | 9.23 |
|  | Median | - | 3.0 | 2.0 | 2.0 | 2.0 |
|  | Minimum | - | 2 | 1 | 1 | 1 |
|  | Maximum | - | 33 | 3 | 33 | 33 |

| Preferred Term (unit)  Most Recent Dose | Statistic | Pooled Placebo (N=10) n (%) | 20 ug ReCOV (N=20) n (%) | 40 ug ReCOV (N=19) n (%) | Pooled ReCOV (N=39) n (%) | Overall (N=49) n (%) |
| --- | --- | --- | --- | --- | --- | --- |
| Headache (days) | | | | | | |
| First Dose | n | 1 | 0 | 4 | 4 | 5 |
|  | Mean | 2.0 | - | 3.8 | 3.8 | 3.4 |
|  | SD | - | - | 0.96 | 0.96 | 1.14 |
|  | Median | 2.0 | - | 3.5 | 3.5 | 3.0 |
|  | Minimum | 2 | - | 3 | 3 | 2 |
|  | Maximum | 2 | - | 5 | 5 | 5 |
| Second Dose | n | 2 | 4 | 5 | 9 | 11 |
|  | Mean | 1.0 | 1.8 | 2.6 | 2.2 | 2.0 |
|  | SD | 0.00 | 0.50 | 1.34 | 1.09 | 1.10 |
|  | Median | 1.0 | 2.0 | 2.0 | 2.0 | 2.0 |
|  | Minimum | 1 | 1 | 2 | 1 | 1 |
|  | Maximum | 1 | 2 | 5 | 5 | 5 |
| Pyrexia (days) | | | | | | |
| Second Dose | n | 0 | 2 | 4 | 6 | 6 |
|  | Mean | - | 3.0 | 2.3 | 2.5 | 2.5 |
|  | SD | - | 1.41 | 0.50 | 0.84 | 0.84 |
|  | Median | - | 3.0 | 2.0 | 2.0 | 2.0 |
|  | Minimum | - | 2 | 2 | 2 | 2 |
|  | Maximum | - | 4 | 3 | 4 | 4 |

| Preferred Term (unit)  Most Recent Dose | Statistic | Pooled Placebo (N=10) n (%) | 20 ug ReCOV (N=20) n (%) | 40 ug ReCOV (N=19) n (%) | Pooled ReCOV (N=39) n (%) | Overall (N=49) n (%) |
| --- | --- | --- | --- | --- | --- | --- |
| Nausea (days) | | | | | | |
| First Dose | n | 1 | 0 | 1 | 1 | 2 |
|  | Mean | 3.0 | - | 3.0 | 3.0 | 3.0 |
|  | SD | - | - | - | - | 0.00 |
|  | Median | 3.0 | - | 3.0 | 3.0 | 3.0 |
|  | Minimum | 3 | - | 3 | 3 | 3 |
|  | Maximum | 3 | - | 3 | 3 | 3 |
| Second Dose | n | 0 | 0 | 2 | 2 | 2 |
|  | Mean | - | - | 1.5 | 1.5 | 1.5 |
|  | SD | - | - | 0.71 | 0.71 | 0.71 |
|  | Median | - | - | 1.5 | 1.5 | 1.5 |
|  | Minimum | - | - | 1 | 1 | 1 |
|  | Maximum | - | - | 2 | 2 | 2 |
| Diarrhoea (days) | | | | | | |
| First Dose | n | 0 | 0 | 1 | 1 | 1 |
|  | Mean | - | - | 3.0 | 3.0 | 3.0 |
|  | SD | - | - | - | - | - |
|  | Median | - | - | 3.0 | 3.0 | 3.0 |
|  | Minimum | - | - | 3 | 3 | 3 |
|  | Maximum | - | - | 3 | 3 | 3 |

| Preferred Term (unit)  Most Recent Dose | Statistic | Pooled Placebo (N=20) n (%) | 20 ug ReCOV (N=40) n (%) | 40 ug ReCOV (N=39) n (%) | Pooled ReCOV (N=79) n (%) | Overall (N=99) n (%) |
| --- | --- | --- | --- | --- | --- | --- |
| Fatigue (days) | | | | | | |
| First Dose | n | 2 | 5 | 6 | 11 | 13 |
|  | Mean | 20.0 | 3.2 | 9.5 | 6.6 | 8.7 |
|  | SD | 18.38 | 1.79 | 11.64 | 8.94 | 10.95 |
|  | Median | 20.0 | 2.0 | 5.5 | 4.0 | 5.0 |
|  | Minimum | 7 | 2 | 2 | 2 | 2 |
|  | Maximum | 33 | 6 | 33 | 33 | 33 |
| Second Dose | n | 2 | 17 | 10 | 27 | 29 |
|  | Mean | 5.0 | 3.8 | 6.7 | 4.9 | 4.9 |
|  | SD | 1.41 | 4.43 | 9.53 | 6.75 | 6.51 |
|  | Median | 5.0 | 2.0 | 3.0 | 2.0 | 3.0 |
|  | Minimum | 4 | 1 | 2 | 1 | 1 |
|  | Maximum | 6 | 19 | 33 | 33 | 33 |

| Preferred Term (unit)  Most Recent Dose | Statistic | Pooled Placebo (N=20) n (%) | 20 ug ReCOV (N=40) n (%) | 40 ug ReCOV (N=39) n (%) | Pooled ReCOV (N=79) n (%) | Overall (N=99) n (%) |
| --- | --- | --- | --- | --- | --- | --- |
| Headache (days) | | | | | | |
| First Dose | n | 1 | 1 | 6 | 7 | 8 |
|  | Mean | 2.0 | 3.0 | 3.5 | 3.4 | 3.3 |
|  | SD | - | - | 1.52 | 1.40 | 1.39 |
|  | Median | 2.0 | 3.0 | 3.5 | 3.0 | 3.0 |
|  | Minimum | 2 | 3 | 1 | 1 | 1 |
|  | Maximum | 2 | 3 | 5 | 5 | 5 |
| Second Dose | n | 3 | 12 | 10 | 22 | 25 |
|  | Mean | 1.0 | 2.7 | 2.5 | 2.6 | 2.4 |
|  | SD | 0.00 | 2.39 | 0.97 | 1.84 | 1.80 |
|  | Median | 1.0 | 2.0 | 2.0 | 2.0 | 2.0 |
|  | Minimum | 1 | 1 | 2 | 1 | 1 |
|  | Maximum | 1 | 10 | 5 | 10 | 10 |

| Preferred Term (unit)  Most Recent Dose | Statistic | Pooled Placebo (N=20) n (%) | 20 ug ReCOV (N=40) n (%) | 40 ug ReCOV (N=39) n (%) | Pooled ReCOV (N=79) n (%) | Overall (N=99) n (%) |
| --- | --- | --- | --- | --- | --- | --- |
| Myalgia (days) | | | | | | |
| First Dose | n | 1 | 1 | 4 | 5 | 6 |
|  | Mean | 11.0 | 2.0 | 4.8 | 4.2 | 5.3 |
|  | SD | - | - | 1.50 | 1.79 | 3.20 |
|  | Median | 11.0 | 2.0 | 5.0 | 4.0 | 5.0 |
|  | Minimum | 11 | 2 | 3 | 2 | 2 |
|  | Maximum | 11 | 2 | 6 | 6 | 11 |
| Second Dose | n | 0 | 14 | 10 | 24 | 24 |
|  | Mean | - | 5.5 | 2.4 | 4.2 | 4.2 |
|  | SD | - | 8.16 | 0.84 | 6.35 | 6.35 |
|  | Median | - | 3.0 | 2.0 | 3.0 | 3.0 |
|  | Minimum | - | 1 | 1 | 1 | 1 |
|  | Maximum | - | 33 | 4 | 33 | 33 |
| Pyrexia (days) | | | | | | |
| Second Dose | n | 0 | 6 | 4 | 10 | 10 |
|  | Mean | - | 3.3 | 2.3 | 2.9 | 2.9 |
|  | SD | - | 2.07 | 0.50 | 1.66 | 1.66 |
|  | Median | - | 3.0 | 2.0 | 2.5 | 2.5 |
|  | Minimum | - | 1 | 2 | 1 | 1 |
|  | Maximum | - | 7 | 3 | 7 | 7 |

| Preferred Term (unit)  Most Recent Dose | Statistic | Pooled Placebo (N=20) n (%) | 20 ug ReCOV (N=40) n (%) | 40 ug ReCOV (N=39) n (%) | Pooled ReCOV (N=79) n (%) | Overall (N=99) n (%) |
| --- | --- | --- | --- | --- | --- | --- |
| Nausea (days) | | | | | | |
| First Dose | n | 1 | 0 | 1 | 1 | 2 |
|  | Mean | 3.0 | - | 3.0 | 3.0 | 3.0 |
|  | SD | - | - | - | - | 0.00 |
|  | Median | 3.0 | - | 3.0 | 3.0 | 3.0 |
|  | Minimum | 3 | - | 3 | 3 | 3 |
|  | Maximum | 3 | - | 3 | 3 | 3 |
| Second Dose | n | 0 | 2 | 5 | 7 | 7 |
|  | Mean | - | 3.5 | 2.2 | 2.6 | 2.6 |
|  | SD | - | 3.54 | 1.10 | 1.81 | 1.81 |
|  | Median | - | 3.5 | 2.0 | 2.0 | 2.0 |
|  | Minimum | - | 1 | 1 | 1 | 1 |
|  | Maximum | - | 6 | 4 | 6 | 6 |

| Preferred Term (unit)  Most Recent Dose | Statistic | Pooled Placebo (N=20) n (%) | 20 ug ReCOV (N=40) n (%) | 40 ug ReCOV (N=39) n (%) | Pooled ReCOV (N=79) n (%) | Overall (N=99) n (%) |
| --- | --- | --- | --- | --- | --- | --- |
| Diarrhoea (days) | | | | | | |
| First Dose | n | 0 | 0 | 1 | 1 | 1 |
|  | Mean | - | - | 3.0 | 3.0 | 3.0 |
|  | SD | - | - | - | - | - |
|  | Median | - | - | 3.0 | 3.0 | 3.0 |
|  | Minimum | - | - | 3 | 3 | 3 |
|  | Maximum | - | - | 3 | 3 | 3 |
| Second Dose | n | 1 | 0 | 0 | 0 | 1 |
|  | Mean | 2.0 | - | - | - | 2.0 |
|  | SD | - | - | - | - | - |
|  | Median | 2.0 | - | - | - | 2.0 |
|  | Minimum | 2 | - | - | - | 2 |
|  | Maximum | 2 | - | - | - | 2 |
| Vomiting (days) | | | | | | |
| Second Dose | n | 0 | 1 | 0 | 1 | 1 |
|  | Mean | - | 3.0 | - | 3.0 | 3.0 |
|  | SD | - | - | - | - | - |
|  | Median | - | 3.0 | - | 3.0 | 3.0 |
|  | Minimum | - | 3 | - | 3 | 3 |
|  | Maximum | - | 3 | - | 3 | 3 |
