## Supplemental Table 6 for "The preliminary safety and immunogenicity results of a randomized, double-blind, placebo-controlled Phase I trial for a recombinant two-component subunit SARS-CoV-2 vaccine ReCOV"

| System Organ Class  Preferred Term | Severity | Pooled Placebo (N=10) n (%) | 20 ug ReCOV (N=20) n (%) | 40 ug ReCOV (N=20) n (%) | Pooled ReCOV (N=40) n (%) | Overall (N=50) n (%) |
| --- | --- | --- | --- | --- | --- | --- |
| At least one Unsolicited TEAE | Mild (Grade 1) | 2 (20.0) | 13 (65.0) | 10 (50.0) | 23 (57.5) | 25 (50.0) |
|  | Moderate (Grade 2) | 0 | 1 (5.0) | 2 (10.0) | 3 (7.5) | 3 (6.0) |
|  | Severe (Grade 3) | 0 | 0 | 0 | 0 | 0 |
|  | Potentially life-threatening (Grade 4) | 0 | 0 | 0 | 0 | 0 |
| General disorders and administration site conditions | Mild (Grade 1) | 0 | 6 (30.0) | 5 (25.0) | 11 (27.5) | 11 (22.0) |
|  | Moderate (Grade 2) | 0 | 0 | 2 (10.0) | 2 (5.0) | 2 (4.0) |
| Influenza like illness | Mild (Grade 1) | 0 | 1 (5.0) | 2 (10.0) | 3 (7.5) | 3 (6.0) |
|  | Moderate (Grade 2) | 0 | 0 | 1 (5.0) | 1 (2.5) | 1 (2.0) |
| Chest pain | Mild (Grade 1) | 0 | 0 | 2 (10.0) | 2 (5.0) | 2 (4.0) |
| Chills | Mild (Grade 1) | 0 | 1 (5.0) | 1 (5.0) | 2 (5.0) | 2 (4.0) |
| Fatigue | Mild (Grade 1) | 0 | 1 (5.0) | 1 (5.0) | 2 (5.0) | 2 (4.0) |
| Injection site pruritus | Mild (Grade 1) | 0 | 1 (5.0) | 1 (5.0) | 2 (5.0) | 2 (4.0) |
| Injection site erythema | Mild (Grade 1) | 0 | 0 | 1 (5.0) | 1 (2.5) | 1 (2.0) |

| System Organ Class  Preferred Term | Severity | Pooled Placebo (N=10) n (%) | 20 ug ReCOV (N=20) n (%) | 40 ug ReCOV (N=20) n (%) | Pooled ReCOV (N=40) n (%) | Overall (N=50) n (%) |
| --- | --- | --- | --- | --- | --- | --- |
| General disorders and administration site conditions (continued) |  |  |  |  |  |  |
| Injection site rash | Moderate (Grade 2) | 0 | 0 | 1 (5.0) | 1 (2.5) | 1 (2.0) |
| Injection site reaction | Mild (Grade 1) | 0 | 1 (5.0) | 0 | 1 (2.5) | 1 (2.0) |
| Injection site swelling | Mild (Grade 1) | 0 | 0 | 1 (5.0) | 1 (2.5) | 1 (2.0) |
| Reactogenicity event | Mild (Grade 1) | 0 | 1 (5.0) | 0 | 1 (2.5) | 1 (2.0) |
| Gastrointestinal disorders | Mild (Grade 1) | 0 | 1 (5.0) | 6 (30.0) | 7 (17.5) | 7 (14.0) |
|  | Moderate (Grade 2) | 0 | 1 (5.0) | 0 | 1 (2.5) | 1 (2.0) |
| Diarrhoea | Mild (Grade 1) | 0 | 0 | 2 (10.0) | 2 (5.0) | 2 (4.0) |
| Abdominal discomfort | Mild (Grade 1) | 0 | 0 | 1 (5.0) | 1 (2.5) | 1 (2.0) |
| Abdominal pain | Mild (Grade 1) | 0 | 0 | 1 (5.0) | 1 (2.5) | 1 (2.0) |
| Dyspepsia | Mild (Grade 1) | 0 | 0 | 1 (5.0) | 1 (2.5) | 1 (2.0) |
| Gastrooesophageal reflux disease | Mild (Grade 1) | 0 | 0 | 1 (5.0) | 1 (2.5) | 1 (2.0) |
| Lip dry | Mild (Grade 1) | 0 | 1 (5.0) | 0 | 1 (2.5) | 1 (2.0) |

| System Organ Class  Preferred Term | Severity | Pooled Placebo (N=10) n (%) | 20 ug ReCOV (N=20) n (%) | 40 ug ReCOV (N=20) n (%) | Pooled ReCOV (N=40) n (%) | Overall (N=50) n (%) |
| --- | --- | --- | --- | --- | --- | --- |
| Gastrointestinal disorders (continued) |  |  |  |  |  |  |
| Paraesthesia oral | Mild (Grade 1) | 0 | 0 | 1 (5.0) | 1 (2.5) | 1 (2.0) |
| Toothache | Moderate (Grade 2) | 0 | 1 (5.0) | 0 | 1 (2.5) | 1 (2.0) |
| Infections and infestations | Mild (Grade 1) | 1 (10.0) | 4 (20.0) | 3 (15.0) | 7 (17.5) | 8 (16.0) |
| Upper respiratory tract infection | Mild (Grade 1) | 1 (10.0) | 4 (20.0) | 2 (10.0) | 6 (15.0) | 7 (14.0) |
| Gastrointestinal viral infection | Mild (Grade 1) | 0 | 0 | 1 (5.0) | 1 (2.5) | 1 (2.0) |
| Pharyngitis | Mild (Grade 1) | 0 | 0 | 1 (5.0) | 1 (2.5) | 1 (2.0) |
| Urinary tract infection | Mild (Grade 1) | 0 | 1 (5.0) | 0 | 1 (2.5) | 1 (2.0) |
| Respiratory, thoracic and mediastinal disorders | Mild (Grade 1) | 1 (10.0) | 3 (15.0) | 1 (5.0) | 4 (10.0) | 5 (10.0) |
| Rhinorrhoea | Mild (Grade 1) | 1 (10.0) | 1 (5.0) | 0 | 1 (2.5) | 2 (4.0) |
| Nasal congestion | Mild (Grade 1) | 0 | 1 (5.0) | 0 | 1 (2.5) | 1 (2.0) |
| Oropharyngeal pain | Mild (Grade 1) | 0 | 1 (5.0) | 0 | 1 (2.5) | 1 (2.0) |

| System Organ Class  Preferred Term | Severity | Pooled Placebo (N=10) n (%) | 20 ug ReCOV (N=20) n (%) | 40 ug ReCOV (N=20) n (%) | Pooled ReCOV (N=40) n (%) | Overall (N=50) n (%) |
| --- | --- | --- | --- | --- | --- | --- |
| Respiratory, thoracic and mediastinal disorders (continued) |  |  |  |  |  |  |
| Productive cough | Mild (Grade 1) | 0 | 0 | 1 (5.0) | 1 (2.5) | 1 (2.0) |
| Nervous system disorders | Mild (Grade 1) | 0 | 3 (15.0) | 1 (5.0) | 4 (10.0) | 4 (8.0) |
| Headache | Mild (Grade 1) | 0 | 2 (10.0) | 1 (5.0) | 3 (7.5) | 3 (6.0) |
| Ageusia | Mild (Grade 1) | 0 | 1 (5.0) | 0 | 1 (2.5) | 1 (2.0) |
| Dizziness | Mild (Grade 1) | 0 | 0 | 1 (5.0) | 1 (2.5) | 1 (2.0) |
| Tension headache | Mild (Grade 1) | 0 | 1 (5.0) | 0 | 1 (2.5) | 1 (2.0) |
| Skin and subcutaneous tissue disorders | Mild (Grade 1) | 0 | 0 | 2 (10.0) | 2 (5.0) | 2 (4.0) |
| Erythema | Mild (Grade 1) | 0 | 0 | 1 (5.0) | 1 (2.5) | 1 (2.0) |
| Pruritus | Mild (Grade 1) | 0 | 0 | 1 (5.0) | 1 (2.5) | 1 (2.0) |
| Rash | Mild (Grade 1) | 0 | 0 | 1 (5.0) | 1 (2.5) | 1 (2.0) |

| System Organ Class  Preferred Term | Severity | Pooled Placebo (N=10) n (%) | 20 ug ReCOV (N=20) n (%) | 40 ug ReCOV (N=20) n (%) | Pooled ReCOV (N=40) n (%) | Overall (N=50) n (%) |
| --- | --- | --- | --- | --- | --- | --- |
| Cardiac disorders | Mild (Grade 1) | 0 | 1 (5.0) | 0 | 1 (2.5) | 1 (2.0) |
| Palpitations | Mild (Grade 1) | 0 | 1 (5.0) | 0 | 1 (2.5) | 1 (2.0) |
| Injury, poisoning and procedural complications | Mild (Grade 1) | 0 | 0 | 1 (5.0) | 1 (2.5) | 1 (2.0) |
| Muscle strain | Mild (Grade 1) | 0 | 0 | 1 (5.0) | 1 (2.5) | 1 (2.0) |
| Metabolism and nutrition disorders | Mild (Grade 1) | 0 | 1 (5.0) | 0 | 1 (2.5) | 1 (2.0) |
| Decreased appetite | Mild (Grade 1) | 0 | 1 (5.0) | 0 | 1 (2.5) | 1 (2.0) |
| Musculoskeletal and connective tissue disorders | Mild (Grade 1) | 0 | 0 | 1 (5.0) | 1 (2.5) | 1 (2.0) |
| Musculoskeletal chest pain | Mild (Grade 1) | 0 | 0 | 1 (5.0) | 1 (2.5) | 1 (2.0) |
| Reproductive system and breast disorders | Mild (Grade 1) | 0 | 0 | 1 (5.0) | 1 (2.5) | 1 (2.0) |
| Breast tenderness | Mild (Grade 1) | 0 | 0 | 1 (5.0) | 1 (2.5) | 1 (2.0) |

| System Organ Class  Preferred Term | Severity | Pooled Placebo (N=10) n (%) | 20 ug ReCOV (N=20) n (%) | 40 ug ReCOV (N=20) n (%) | Pooled ReCOV (N=40) n (%) | Overall (N=50) n (%) |
| --- | --- | --- | --- | --- | --- | --- |
| Surgical and medical procedures | Mild (Grade 1) | 0 | 0 | 1 (5.0) | 1 (2.5) | 1 (2.0) |
| Tooth repair | Mild (Grade 1) | 0 | 0 | 1 (5.0) | 1 (2.5) | 1 (2.0) |

| System Organ Class  Preferred Term | Severity | Pooled Placebo (N=10) n (%) | 20 ug ReCOV (N=20) n (%) | 40 ug ReCOV (N=19) n (%) | Pooled ReCOV (N=39) n (%) | Overall (N=49) n (%) |
| --- | --- | --- | --- | --- | --- | --- |
| At least one Unsolicited TEAE | Mild (Grade 1) | 5 (50.0) | 11 (55.0) | 12 (63.2) | 23 (59.0) | 28 (57.1) |
|  | Moderate (Grade 2) | 2 (20.0) | 0 | 0 | 0 | 2 (4.1) |
|  | Severe (Grade 3) | 0 | 0 | 0 | 0 | 0 |
|  | Potentially life-threatening (Grade 4) | 0 | 0 | 0 | 0 | 0 |
| General disorders and administration site conditions | Mild (Grade 1) | 3 (30.0) | 2 (10.0) | 7 (36.8) | 9 (23.1) | 12 (24.5) |
| Injection site pruritus | Mild (Grade 1) | 1 (10.0) | 0 | 2 (10.5) | 2 (5.1) | 3 (6.1) |
| Fatigue | Mild (Grade 1) | 0 | 0 | 2 (10.5) | 2 (5.1) | 2 (4.1) |
| Influenza like illness | Mild (Grade 1) | 0 | 1 (5.0) | 1 (5.3) | 2 (5.1) | 2 (4.1) |
| Chest pain | Mild (Grade 1) | 1 (10.0) | 0 | 0 | 0 | 1 (2.0) |
| Injection site discomfort | Mild (Grade 1) | 0 | 0 | 1 (5.3) | 1 (2.6) | 1 (2.0) |
| Injection site irritation | Mild (Grade 1) | 0 | 1 (5.0) | 0 | 1 (2.6) | 1 (2.0) |
| Injection site pain | Mild (Grade 1) | 0 | 0 | 1 (5.3) | 1 (2.6) | 1 (2.0) |

| System Organ Class  Preferred Term | Severity | Pooled Placebo (N=10) n (%) | 20 ug ReCOV (N=20) n (%) | 40 ug ReCOV (N=19) n (%) | Pooled ReCOV (N=39) n (%) | Overall (N=49) n (%) |
| --- | --- | --- | --- | --- | --- | --- |
| General disorders and administration site conditions (continued) |  |  |  |  |  |  |
| Injection site swelling | Mild (Grade 1) | 0 | 0 | 1 (5.3) | 1 (2.6) | 1 (2.0) |
| Non-cardiac chest pain | Mild (Grade 1) | 1 (10.0) | 0 | 0 | 0 | 1 (2.0) |
| Vessel puncture site bruise | Mild (Grade 1) | 1 (10.0) | 0 | 0 | 0 | 1 (2.0) |
| Infections and infestations | Mild (Grade 1) | 4 (40.0) | 3 (15.0) | 3 (15.8) | 6 (15.4) | 10 (20.4) |
| Upper respiratory tract infection | Mild (Grade 1) | 3 (30.0) | 2 (10.0) | 1 (5.3) | 3 (7.7) | 6 (12.2) |
| Conjunctivitis | Mild (Grade 1) | 0 | 1 (5.0) | 0 | 1 (2.6) | 1 (2.0) |
| Infected dermal cyst | Mild (Grade 1) | 1 (10.0) | 0 | 0 | 0 | 1 (2.0) |
| Otitis externa | Mild (Grade 1) | 0 | 0 | 1 (5.3) | 1 (2.6) | 1 (2.0) |
| Sinusitis | Mild (Grade 1) | 0 | 0 | 1 (5.3) | 1 (2.6) | 1 (2.0) |
| Musculoskeletal and connective tissue disorders | Mild (Grade 1) | 2 (20.0) | 4 (20.0) | 1 (5.3) | 5 (12.8) | 7 (14.3) |
| Back pain | Mild (Grade 1) | 1 (10.0) | 1 (5.0) | 0 | 1 (2.6) | 2 (4.1) |

| System Organ Class  Preferred Term | Severity | Pooled Placebo (N=10) n (%) | 20 ug ReCOV (N=20) n (%) | 40 ug ReCOV (N=19) n (%) | Pooled ReCOV (N=39) n (%) | Overall (N=49) n (%) |
| --- | --- | --- | --- | --- | --- | --- |
| Musculoskeletal and connective tissue disorders (continued) |  |  |  |  |  |  |
| Arthralgia | Mild (Grade 1) | 0 | 1 (5.0) | 0 | 1 (2.6) | 1 (2.0) |
| Musculoskeletal pain | Mild (Grade 1) | 0 | 0 | 1 (5.3) | 1 (2.6) | 1 (2.0) |
| Musculoskeletal stiffness | Mild (Grade 1) | 0 | 1 (5.0) | 0 | 1 (2.6) | 1 (2.0) |
| Myalgia | Mild (Grade 1) | 0 | 1 (5.0) | 0 | 1 (2.6) | 1 (2.0) |
| Pain in extremity | Mild (Grade 1) | 1 (10.0) | 0 | 0 | 0 | 1 (2.0) |
| Nervous system disorders | Mild (Grade 1) | 1 (10.0) | 2 (10.0) | 4 (21.1) | 6 (15.4) | 7 (14.3) |
| Dizziness | Mild (Grade 1) | 1 (10.0) | 2 (10.0) | 2 (10.5) | 4 (10.3) | 5 (10.2) |
| Headache | Mild (Grade 1) | 0 | 0 | 2 (10.5) | 2 (5.1) | 2 (4.1) |
| Eye disorders | Mild (Grade 1) | 1 (10.0) | 3 (15.0) | 1 (5.3) | 4 (10.3) | 5 (10.2) |
| Blepharospasm | Mild (Grade 1) | 0 | 2 (10.0) | 0 | 2 (5.1) | 2 (4.1) |
| Lacrimation increased | Mild (Grade 1) | 1 (10.0) | 0 | 1 (5.3) | 1 (2.6) | 2 (4.1) |

| System Organ Class  Preferred Term | Severity | Pooled Placebo (N=10) n (%) | 20 ug ReCOV (N=20) n (%) | 40 ug ReCOV (N=19) n (%) | Pooled ReCOV (N=39) n (%) | Overall (N=49) n (%) |
| --- | --- | --- | --- | --- | --- | --- |
| Eye disorders (continued) |  |  |  |  |  |  |
| Periorbital pain | Mild (Grade 1) | 0 | 1 (5.0) | 0 | 1 (2.6) | 1 (2.0) |
| Injury, poisoning and procedural complications | Mild (Grade 1) | 2 (20.0) | 2 (10.0) | 1 (5.3) | 3 (7.7) | 5 (10.2) |
| Contusion | Mild (Grade 1) | 1 (10.0) | 0 | 1 (5.3) | 1 (2.6) | 2 (4.1) |
| Animal bite | Mild (Grade 1) | 0 | 1 (5.0) | 0 | 1 (2.6) | 1 (2.0) |
| Animal scratch | Mild (Grade 1) | 0 | 1 (5.0) | 0 | 1 (2.6) | 1 (2.0) |
| Arthropod bite | Mild (Grade 1) | 1 (10.0) | 0 | 0 | 0 | 1 (2.0) |
| Skin laceration | Mild (Grade 1) | 0 | 1 (5.0) | 0 | 1 (2.6) | 1 (2.0) |
| Skin and subcutaneous tissue disorders | Mild (Grade 1) | 0 | 3 (15.0) | 0 | 3 (7.7) | 3 (6.1) |
|  | Moderate (Grade 2) | 1 (10.0) | 0 | 0 | 0 | 1 (2.0) |
| Rash | Mild (Grade 1) | 0 | 1 (5.0) | 0 | 1 (2.6) | 1 (2.0) |
|  | Moderate (Grade 2) | 1 (10.0) | 0 | 0 | 0 | 1 (2.0) |
| Blister | Mild (Grade 1) | 0 | 1 (5.0) | 0 | 1 (2.6) | 1 (2.0) |

| System Organ Class  Preferred Term | Severity | Pooled Placebo (N=10) n (%) | 20 ug ReCOV (N=20) n (%) | 40 ug ReCOV (N=19) n (%) | Pooled ReCOV (N=39) n (%) | Overall (N=49) n (%) |
| --- | --- | --- | --- | --- | --- | --- |
| Skin and subcutaneous tissue disorders (continued) |  |  |  |  |  |  |
| Pruritus | Mild (Grade 1) | 0 | 1 (5.0) | 0 | 1 (2.6) | 1 (2.0) |
| Gastrointestinal disorders | Mild (Grade 1) | 0 | 1 (5.0) | 0 | 1 (2.6) | 1 (2.0) |
|  | Moderate (Grade 2) | 1 (10.0) | 0 | 0 | 0 | 1 (2.0) |
| Abdominal pain | Mild (Grade 1) | 1 (10.0) | 0 | 0 | 0 | 1 (2.0) |
| Eructation | Mild (Grade 1) | 0 | 1 (5.0) | 0 | 1 (2.6) | 1 (2.0) |
| Toothache | Moderate (Grade 2) | 1 (10.0) | 0 | 0 | 0 | 1 (2.0) |
| Respiratory, thoracic and mediastinal disorders | Mild (Grade 1) | 0 | 0 | 1 (5.3) | 1 (2.6) | 1 (2.0) |
|  | Moderate (Grade 2) | 1 (10.0) | 0 | 0 | 0 | 1 (2.0) |
| Rhinorrhoea | Mild (Grade 1) | 1 (10.0) | 0 | 1 (5.3) | 1 (2.6) | 2 (4.1) |
| Rhinalgia | Moderate (Grade 2) | 1 (10.0) | 0 | 0 | 0 | 1 (2.0) |
| Throat irritation | Mild (Grade 1) | 1 (10.0) | 0 | 0 | 0 | 1 (2.0) |

| System Organ Class  Preferred Term | Severity | Pooled Placebo (N=10) n (%) | 20 ug ReCOV (N=20) n (%) | 40 ug ReCOV (N=19) n (%) | Pooled ReCOV (N=39) n (%) | Overall (N=49) n (%) |
| --- | --- | --- | --- | --- | --- | --- |
| Metabolism and nutrition disorders | Mild (Grade 1) | 0 | 1 (5.0) | 0 | 1 (2.6) | 1 (2.0) |
| Decreased appetite | Mild (Grade 1) | 0 | 1 (5.0) | 0 | 1 (2.6) | 1 (2.0) |

| System Organ Class  Preferred Term | Severity | Pooled Placebo (N=20) n (%) | 20 ug ReCOV (N=40) n (%) | 40 ug ReCOV (N=39) n (%) | Pooled ReCOV (N=79) n (%) | Overall (N=99) n (%) |
| --- | --- | --- | --- | --- | --- | --- |
| At least one Unsolicited TEAE | Mild (Grade 1) | 7 (35.0) | 24 (60.0) | 22 (56.4) | 46 (58.2) | 53 (53.5) |
|  | Moderate (Grade 2) | 2 (10.0) | 1 (2.5) | 2 (5.1) | 3 (3.8) | 5 (5.1) |
|  | Severe (Grade 3) | 0 | 0 | 0 | 0 | 0 |
|  | Potentially life-threatening (Grade 4) | 0 | 0 | 0 | 0 | 0 |
| General disorders and administration site conditions | Mild (Grade 1) | 3 (15.0) | 8 (20.0) | 12 (30.8) | 20 (25.3) | 23 (23.2) |
|  | Moderate (Grade 2) | 0 | 0 | 2 (5.1) | 2 (2.5) | 2 (2.0) |
| Influenza like illness | Mild (Grade 1) | 0 | 2 (5.0) | 3 (7.7) | 5 (6.3) | 5 (5.1) |
|  | Moderate (Grade 2) | 0 | 0 | 1 (2.6) | 1 (1.3) | 1 (1.0) |
| Injection site pruritus | Mild (Grade 1) | 1 (5.0) | 1 (2.5) | 3 (7.7) | 4 (5.1) | 5 (5.1) |
| Fatigue | Mild (Grade 1) | 0 | 1 (2.5) | 3 (7.7) | 4 (5.1) | 4 (4.0) |
| Chest pain | Mild (Grade 1) | 1 (5.0) | 0 | 2 (5.1) | 2 (2.5) | 3 (3.0) |
| Chills | Mild (Grade 1) | 0 | 1 (2.5) | 1 (2.6) | 2 (2.5) | 2 (2.0) |
| Injection site swelling | Mild (Grade 1) | 0 | 0 | 2 (5.1) | 2 (2.5) | 2 (2.0) |

| System Organ Class  Preferred Term | Severity | Pooled Placebo (N=20) n (%) | 20 ug ReCOV (N=40) n (%) | 40 ug ReCOV (N=39) n (%) | Pooled ReCOV (N=79) n (%) | Overall (N=99) n (%) |
| --- | --- | --- | --- | --- | --- | --- |
| General disorders and administration site conditions (continued) |  |  |  |  |  |  |
| Injection site discomfort | Mild (Grade 1) | 0 | 0 | 1 (2.6) | 1 (1.3) | 1 (1.0) |
| Injection site erythema | Mild (Grade 1) | 0 | 0 | 1 (2.6) | 1 (1.3) | 1 (1.0) |
| Injection site irritation | Mild (Grade 1) | 0 | 1 (2.5) | 0 | 1 (1.3) | 1 (1.0) |
| Injection site pain | Mild (Grade 1) | 0 | 0 | 1 (2.6) | 1 (1.3) | 1 (1.0) |
| Injection site rash | Moderate (Grade 2) | 0 | 0 | 1 (2.6) | 1 (1.3) | 1 (1.0) |
| Injection site reaction | Mild (Grade 1) | 0 | 1 (2.5) | 0 | 1 (1.3) | 1 (1.0) |
| Non-cardiac chest pain | Mild (Grade 1) | 1 (5.0) | 0 | 0 | 0 | 1 (1.0) |
| Reactogenicity event | Mild (Grade 1) | 0 | 1 (2.5) | 0 | 1 (1.3) | 1 (1.0) |
| Vessel puncture site bruise | Mild (Grade 1) | 1 (5.0) | 0 | 0 | 0 | 1 (1.0) |
| Infections and infestations | Mild (Grade 1) | 5 (25.0) | 7 (17.5) | 6 (15.4) | 13 (16.5) | 18 (18.2) |
| Upper respiratory tract infection | Mild (Grade 1) | 4 (20.0) | 6 (15.0) | 3 (7.7) | 9 (11.4) | 13 (13.1) |

| System Organ Class  Preferred Term | Severity | Pooled Placebo (N=20) n (%) | 20 ug ReCOV (N=40) n (%) | 40 ug ReCOV (N=39) n (%) | Pooled ReCOV (N=79) n (%) | Overall (N=99) n (%) |
| --- | --- | --- | --- | --- | --- | --- |
| Infections and infestations (continued) |  |  |  |  |  |  |
| Conjunctivitis | Mild (Grade 1) | 0 | 1 (2.5) | 0 | 1 (1.3) | 1 (1.0) |
| Gastrointestinal viral infection | Mild (Grade 1) | 0 | 0 | 1 (2.6) | 1 (1.3) | 1 (1.0) |
| Infected dermal cyst | Mild (Grade 1) | 1 (5.0) | 0 | 0 | 0 | 1 (1.0) |
| Otitis externa | Mild (Grade 1) | 0 | 0 | 1 (2.6) | 1 (1.3) | 1 (1.0) |
| Pharyngitis | Mild (Grade 1) | 0 | 0 | 1 (2.6) | 1 (1.3) | 1 (1.0) |
| Sinusitis | Mild (Grade 1) | 0 | 0 | 1 (2.6) | 1 (1.3) | 1 (1.0) |
| Urinary tract infection | Mild (Grade 1) | 0 | 1 (2.5) | 0 | 1 (1.3) | 1 (1.0) |
| Nervous system disorders | Mild (Grade 1) | 1 (5.0) | 5 (12.5) | 5 (12.8) | 10 (12.7) | 11 (11.1) |
| Dizziness | Mild (Grade 1) | 1 (5.0) | 2 (5.0) | 3 (7.7) | 5 (6.3) | 6 (6.1) |
| Headache | Mild (Grade 1) | 0 | 2 (5.0) | 3 (7.7) | 5 (6.3) | 5 (5.1) |
| Ageusia | Mild (Grade 1) | 0 | 1 (2.5) | 0 | 1 (1.3) | 1 (1.0) |

| System Organ Class  Preferred Term | Severity | Pooled Placebo (N=20) n (%) | 20 ug ReCOV (N=40) n (%) | 40 ug ReCOV (N=39) n (%) | Pooled ReCOV (N=79) n (%) | Overall (N=99) n (%) |
| --- | --- | --- | --- | --- | --- | --- |
| Nervous system disorders (continued) |  |  |  |  |  |  |
| Tension headache | Mild (Grade 1) | 0 | 1 (2.5) | 0 | 1 (1.3) | 1 (1.0) |
| Gastrointestinal disorders | Mild (Grade 1) | 0 | 2 (5.0) | 6 (15.4) | 8 (10.1) | 8 (8.1) |
|  | Moderate (Grade 2) | 1 (5.0) | 1 (2.5) | 0 | 1 (1.3) | 2 (2.0) |
| Abdominal pain | Mild (Grade 1) | 1 (5.0) | 0 | 1 (2.6) | 1 (1.3) | 2 (2.0) |
| Diarrhoea | Mild (Grade 1) | 0 | 0 | 2 (5.1) | 2 (2.5) | 2 (2.0) |
| Toothache | Moderate (Grade 2) | 1 (5.0) | 1 (2.5) | 0 | 1 (1.3) | 2 (2.0) |
| Abdominal discomfort | Mild (Grade 1) | 0 | 0 | 1 (2.6) | 1 (1.3) | 1 (1.0) |
| Dyspepsia | Mild (Grade 1) | 0 | 0 | 1 (2.6) | 1 (1.3) | 1 (1.0) |
| Eructation | Mild (Grade 1) | 0 | 1 (2.5) | 0 | 1 (1.3) | 1 (1.0) |
| Gastrooesophageal reflux disease | Mild (Grade 1) | 0 | 0 | 1 (2.6) | 1 (1.3) | 1 (1.0) |
| Lip dry | Mild (Grade 1) | 0 | 1 (2.5) | 0 | 1 (1.3) | 1 (1.0) |
| Paraesthesia oral | Mild (Grade 1) | 0 | 0 | 1 (2.6) | 1 (1.3) | 1 (1.0) |

| System Organ Class  Preferred Term | Severity | Pooled Placebo (N=20) n (%) | 20 ug ReCOV (N=40) n (%) | 40 ug ReCOV (N=39) n (%) | Pooled ReCOV (N=79) n (%) | Overall (N=99) n (%) |
| --- | --- | --- | --- | --- | --- | --- |
| Musculoskeletal and connective tissue disorders | Mild (Grade 1) | 2 (10.0) | 4 (10.0) | 2 (5.1) | 6 (7.6) | 8 (8.1) |
| Back pain | Mild (Grade 1) | 1 (5.0) | 1 (2.5) | 0 | 1 (1.3) | 2 (2.0) |
| Arthralgia | Mild (Grade 1) | 0 | 1 (2.5) | 0 | 1 (1.3) | 1 (1.0) |
| Musculoskeletal chest pain | Mild (Grade 1) | 0 | 0 | 1 (2.6) | 1 (1.3) | 1 (1.0) |
| Musculoskeletal pain | Mild (Grade 1) | 0 | 0 | 1 (2.6) | 1 (1.3) | 1 (1.0) |
| Musculoskeletal stiffness | Mild (Grade 1) | 0 | 1 (2.5) | 0 | 1 (1.3) | 1 (1.0) |
| Myalgia | Mild (Grade 1) | 0 | 1 (2.5) | 0 | 1 (1.3) | 1 (1.0) |
| Pain in extremity | Mild (Grade 1) | 1 (5.0) | 0 | 0 | 0 | 1 (1.0) |
| Respiratory, thoracic and mediastinal disorders | Mild (Grade 1) | 1 (5.0) | 3 (7.5) | 2 (5.1) | 5 (6.3) | 6 (6.1) |
|  | Moderate (Grade 2) | 1 (5.0) | 0 | 0 | 0 | 1 (1.0) |
| Rhinorrhoea | Mild (Grade 1) | 2 (10.0) | 1 (2.5) | 1 (2.6) | 2 (2.5) | 4 (4.0) |

| System Organ Class  Preferred Term | Severity | Pooled Placebo (N=20) n (%) | 20 ug ReCOV (N=40) n (%) | 40 ug ReCOV (N=39) n (%) | Pooled ReCOV (N=79) n (%) | Overall (N=99) n (%) |
| --- | --- | --- | --- | --- | --- | --- |
| Respiratory, thoracic and mediastinal disorders (continued) |  |  |  |  |  |  |
| Nasal congestion | Mild (Grade 1) | 0 | 1 (2.5) | 0 | 1 (1.3) | 1 (1.0) |
| Oropharyngeal pain | Mild (Grade 1) | 0 | 1 (2.5) | 0 | 1 (1.3) | 1 (1.0) |
| Productive cough | Mild (Grade 1) | 0 | 0 | 1 (2.6) | 1 (1.3) | 1 (1.0) |
| Rhinalgia | Moderate (Grade 2) | 1 (5.0) | 0 | 0 | 0 | 1 (1.0) |
| Throat irritation | Mild (Grade 1) | 1 (5.0) | 0 | 0 | 0 | 1 (1.0) |
| Injury, poisoning and procedural complications | Mild (Grade 1) | 2 (10.0) | 2 (5.0) | 2 (5.1) | 4 (5.1) | 6 (6.1) |
| Contusion | Mild (Grade 1) | 1 (5.0) | 0 | 1 (2.6) | 1 (1.3) | 2 (2.0) |
| Animal bite | Mild (Grade 1) | 0 | 1 (2.5) | 0 | 1 (1.3) | 1 (1.0) |
| Animal scratch | Mild (Grade 1) | 0 | 1 (2.5) | 0 | 1 (1.3) | 1 (1.0) |
| Arthropod bite | Mild (Grade 1) | 1 (5.0) | 0 | 0 | 0 | 1 (1.0) |
| Muscle strain | Mild (Grade 1) | 0 | 0 | 1 (2.6) | 1 (1.3) | 1 (1.0) |

| System Organ Class  Preferred Term | Severity | Pooled Placebo (N=20) n (%) | 20 ug ReCOV (N=40) n (%) | 40 ug ReCOV (N=39) n (%) | Pooled ReCOV (N=79) n (%) | Overall (N=99) n (%) |
| --- | --- | --- | --- | --- | --- | --- |
| Injury, poisoning and procedural complications (continued) |  |  |  |  |  |  |
| Skin laceration | Mild (Grade 1) | 0 | 1 (2.5) | 0 | 1 (1.3) | 1 (1.0) |
| Skin and subcutaneous tissue disorders | Mild (Grade 1) | 0 | 3 (7.5) | 2 (5.1) | 5 (6.3) | 5 (5.1) |
|  | Moderate (Grade 2) | 1 (5.0) | 0 | 0 | 0 | 1 (1.0) |
| Rash | Mild (Grade 1) | 0 | 1 (2.5) | 1 (2.6) | 2 (2.5) | 2 (2.0) |
|  | Moderate (Grade 2) | 1 (5.0) | 0 | 0 | 0 | 1 (1.0) |
| Pruritus | Mild (Grade 1) | 0 | 1 (2.5) | 1 (2.6) | 2 (2.5) | 2 (2.0) |
| Blister | Mild (Grade 1) | 0 | 1 (2.5) | 0 | 1 (1.3) | 1 (1.0) |
| Erythema | Mild (Grade 1) | 0 | 0 | 1 (2.6) | 1 (1.3) | 1 (1.0) |
| Eye disorders | Mild (Grade 1) | 1 (5.0) | 3 (7.5) | 1 (2.6) | 4 (5.1) | 5 (5.1) |
| Blepharospasm | Mild (Grade 1) | 0 | 2 (5.0) | 0 | 2 (2.5) | 2 (2.0) |
| Lacrimation increased | Mild (Grade 1) | 1 (5.0) | 0 | 1 (2.6) | 1 (1.3) | 2 (2.0) |
| Periorbital pain | Mild (Grade 1) | 0 | 1 (2.5) | 0 | 1 (1.3) | 1 (1.0) |

| System Organ Class  Preferred Term | Severity | Pooled Placebo (N=20) n (%) | 20 ug ReCOV (N=40) n (%) | 40 ug ReCOV (N=39) n (%) | Pooled ReCOV (N=79) n (%) | Overall (N=99) n (%) |
| --- | --- | --- | --- | --- | --- | --- |
| Metabolism and nutrition disorders | Mild (Grade 1) | 0 | 2 (5.0) | 0 | 2 (2.5) | 2 (2.0) |
| Decreased appetite | Mild (Grade 1) | 0 | 2 (5.0) | 0 | 2 (2.5) | 2 (2.0) |
| Cardiac disorders | Mild (Grade 1) | 0 | 1 (2.5) | 0 | 1 (1.3) | 1 (1.0) |
| Palpitations | Mild (Grade 1) | 0 | 1 (2.5) | 0 | 1 (1.3) | 1 (1.0) |
| Reproductive system and breast disorders | Mild (Grade 1) | 0 | 0 | 1 (2.6) | 1 (1.3) | 1 (1.0) |
| Breast tenderness | Mild (Grade 1) | 0 | 0 | 1 (2.6) | 1 (1.3) | 1 (1.0) |
| Surgical and medical procedures | Mild (Grade 1) | 0 | 0 | 1 (2.6) | 1 (1.3) | 1 (1.0) |
| Tooth repair | Mild (Grade 1) | 0 | 0 | 1 (2.6) | 1 (1.3) | 1 (1.0) |
