## Supplemental Table 7 for "The preliminary safety and immunogenicity results of a randomized, double-blind, placebo-controlled Phase I trial for a recombinant two-component subunit SARS-CoV-2 vaccine ReCOV"

| Analyte (Unit) [LLOQ]  Post-Baseline Visit | Statistic | Pooled Placebo (N=10) | 20 ug ReCOV (N=20) | 40 ug ReCOV (N=20) |
| --- | --- | --- | --- | --- |
| SARS-CoV-2 hCoV-19 Australia (IU/mL) [12.4] |  |  |  |  |
| Day 22 | n (%) | 0 | 19 (95.0) | 20 (100.0) |
|  | [95% CI] | [0.0, 30.8] | [75.1, 99.9] | [83.2, 100.0] |
| Day 36 | n (%) | 0 | 20 (100.0) | 20 (100.0) |
|  | [95% CI] | [0.0, 30.8] | [83.2, 100.0] | [83.2, 100.0] |
| Day 52 | n (%) | 0 | 20 (100.0) | 20 (100.0) |
|  | [95% CI] | [0.0, 30.8] | [83.2, 100.0] | [83.2, 100.0] |
| SARS-CoV-2-hCoV-19 Delta Variant (IU/mL) [12.4] |  |  |  |  |
| Day 36 | n (%) | 0 | 20 (100.0) | 20 (100.0) |
|  | [95% CI] | NA | [83.2, 100.0] | [83.2, 100.0] |
| Day 52 | n (%) | 0 | 20 (100.0) | 20 (100.0) |
|  | [95% CI] | NA | [83.2, 100.0] | [83.2, 100.0] |

| Analyte (Unit) [LLOQ]  Post-Baseline Visit | Statistic | Pooled Placebo (N=10) | 20 ug ReCOV (N=20) | 40 ug ReCOV (N=18) |
| --- | --- | --- | --- | --- |
| SARS-CoV-2 hCoV-19 Australia (IU/mL) [12.4] |  |  |  |  |
| Day 22 | n (%) | 0 | 14 (70.0) | 17 (94.4) |
|  | [95% CI] | [0.0, 30.8] | [45.7, 88.1] | [72.7, 99.9] |
| Day 36 | n (%) | 0 | 20 (100.0) | 18 (100.0) |
|  | [95% CI] | [0.0, 30.8] | [83.2, 100.0] | [81.5, 100.0] |
| Day 52 | n (%) | 0 | 20 (100.0) | 18 (100.0) |
|  | [95% CI] | [0.0, 30.8] | [83.2, 100.0] | [81.5, 100.0] |
| SARS-CoV-2-hCoV-19 Delta Variant (IU/mL) [12.4] |  |  |  |  |
| Day 36 | n (%) | 0 | 18 (90.0) | 16 (88.9) |
|  | [95% CI] | NA | [68.3, 98.8] | [65.3, 98.6] |
| Day 52 | n (%) | 0 | 20 (100.0) | 0 |
|  | [95% CI] | NA | [83.2, 100.0] | NA |

| Analyte (Unit) [LLOQ]  Post-Baseline Visit | Statistic | Pooled Placebo (N=20) | 20 ug ReCOV (N=40) | 40 ug ReCOV (N=38) |
| --- | --- | --- | --- | --- |
| SARS-CoV-2 hCoV-19 Australia (IU/mL) [12.4] |  |  |  |  |
| Day 22 | n (%) | 0 | 33 (82.5) | 37 (97.4) |
|  | [95% CI] | [0.0, 16.8] | [67.2, 92.7] | [86.2, 99.9] |
| Day 36 | n (%) | 0 | 40 (100.0) | 38 (100.0) |
|  | [95% CI] | [0.0, 16.8] | [91.2, 100.0] | [90.7, 100.0] |
| Day 52 | n (%) | 0 | 40 (100.0) | 38 (100.0) |
|  | [95% CI] | [0.0, 16.8] | [91.2, 100.0] | [90.7, 100.0] |
| SARS-CoV-2-hCoV-19 Delta Variant (IU/mL) [12.4] |  |  |  |  |
| Day 36 | n (%) | 0 | 38 (95.0) | 36 (94.7) |
|  | [95% CI] | NA | [83.1, 99.4] | [82.3, 99.4] |
| Day 52 | n (%) | 0 | 40 (100.0) | 20 (100.0) |
|  | [95% CI] | NA | [91.2, 100.0] | [83.2, 100.0] |
