## Supplemental Table 8 for "The preliminary safety and immunogenicity results of a randomized, double-blind, placebo-controlled Phase I trial for a recombinant two-component subunit SARS-CoV-2 vaccine ReCOV"

### Table S8 GMT of Neutralizing Antibody against Delta Variant and hCoV-19 Australian Strains at Each Time Point

| **Visit** | **20μg ReCOV (N=40)** | | | **40μg ReCOV (N=38)** | | |
| --- | --- | --- | --- | --- | --- | --- |
|  | **hCoV-19 Australia** | **Delta Variant** | **Reduction (Fold)** | **hCoV-19 Australia** | **Delta Variant** | **Reduction (Fold)** |
| **Mean (95% CI), IU/mL** | | | | | |  |
| Baseline | 6.31 (6.1, 6.5) | 6.20 (NA) | 0.02 | 6.20 (NA) | 6.20 (NA) | 0 |
| Day 36 | 1358.00 (1038.8, 1775.3) | 111.99 (83.6, 150.0) | 11.12 | 952.40 (706.9, 1283.2) | 93.92 (71.2, 123.9) | 9.14 |
| Day 52 | 943.76 (714.4, 1246.7) | 124.26 (100.8, 153.2) | 6.60 | 649.32 (490.8, 859.0) | 106.32 (82.3, 137.3) | 5.11 |

GMT: Geometric mean titers. CI: Confidence Interval. N=Total number of randomized subjects. Baseline is defined as the last available, non-missing assessment (scheduled or unscheduled) prior to first dose. NA: for all the testing values that were below the LLOQ, a uniform value of 0.5*LLOQ was assigned artificially for the purpose of geometric mean calculation. Accordingly, the anti-log 95%CIs were not displayed as the same values, noted as NA accordingly. Note: For the testing of neutralizing antibody against Delta variant, the serum samples only from 20μg and 40μg ReCOV groups had been tested at the central laboratory.

Date source: Table 14.2.1.2.1 (Age Group: All Subjects (18 - <80 years), Strains: SARS-CoV-2 hCoV-19 Australia and SARS-CoV-2-hCoV-19 Delta Variant)
