## Supplemental Table 9 for "The preliminary safety and immunogenicity results of a randomized, double-blind, placebo-controlled Phase I trial for a recombinant two-component subunit SARS-CoV-2 vaccine ReCOV"

| Analyte (Unit)  Post-Baseline Visit | Statistic | Pooled Placebo (N=10) | 20 ug ReCOV (N=20) | 40 ug ReCOV (N=20) |
| --- | --- | --- | --- | --- |
| SARS-CoV-2 S1 RBD (BAU/mL) |  |  |  |  |
| Day 22 | n (%) | 0 | 20 (100.0) | 20 (100.0) |
|  | [95% CI] | [0.0, 30.8] | [83.2, 100.0] | [83.2, 100.0] |
| Day 36 | n (%) | 0 | 20 (100.0) | 20 (100.0) |
|  | [95% CI] | [0.0, 30.8] | [83.2, 100.0] | [83.2, 100.0] |
| Day 52 | n (%) | 0 | 20 (100.0) | 20 (100.0) |
|  | [95% CI] | [0.0, 30.8] | [83.2, 100.0] | [83.2, 100.0] |
| SARS-CoV-2 S1 NTD (AU/mL) |  |  |  |  |
| Day 22 | n (%) | 0 | 20 (100.0) | 20 (100.0) |
|  | [95% CI] | [0.0, 30.8] | [83.2, 100.0] | [83.2, 100.0] |
| Day 36 | n (%) | 0 | 20 (100.0) | 20 (100.0) |
|  | [95% CI] | [0.0, 30.8] | [83.2, 100.0] | [83.2, 100.0] |
| Day 52 | n (%) | 0 | 20 (100.0) | 20 (100.0) |
|  | [95% CI] | [0.0, 30.8] | [83.2, 100.0] | [83.2, 100.0] |

| Analyte (Unit)  Post-Baseline Visit | Statistic | Pooled Placebo (N=10) | 20 ug ReCOV (N=20) | 40 ug ReCOV (N=18) |
| --- | --- | --- | --- | --- |
| SARS-CoV-2 S1 RBD (BAU/mL) |  |  |  |  |
| Day 22 | n (%) | 1 (10.0) | 20 (100.0) | 18 (100.0) |
|  | [95% CI] | [0.3, 44.5] | [83.2, 100.0] | [81.5, 100.0] |
| Day 36 | n (%) | 0 | 20 (100.0) | 18 (100.0) |
|  | [95% CI] | [0.0, 30.8] | [83.2, 100.0] | [81.5, 100.0] |
| Day 52 | n (%) | 0 | 20 (100.0) | 18 (100.0) |
|  | [95% CI] | [0.0, 30.8] | [83.2, 100.0] | [81.5, 100.0] |
| SARS-CoV-2 S1 NTD (AU/mL) |  |  |  |  |
| Day 22 | n (%) | 1 (10.0) | 20 (100.0) | 18 (100.0) |
|  | [95% CI] | [0.3, 44.5] | [83.2, 100.0] | [81.5, 100.0] |
| Day 36 | n (%) | 0 | 20 (100.0) | 18 (100.0) |
|  | [95% CI] | [0.0, 30.8] | [83.2, 100.0] | [81.5, 100.0] |
| Day 52 | n (%) | 0 | 20 (100.0) | 18 (100.0) |
|  | [95% CI] | [0.0, 30.8] | [83.2, 100.0] | [81.5, 100.0] |

| Analyte (Unit)  Post-Baseline Visit | Statistic | Pooled Placebo (N=20) | 20 ug ReCOV (N=40) | 40 ug ReCOV (N=38) |
| --- | --- | --- | --- | --- |
| SARS-CoV-2 S1 RBD (BAU/mL) |  |  |  |  |
| Day 22 | n (%) | 1 (5.0) | 40 (100.0) | 38 (100.0) |
|  | [95% CI] | [0.1, 24.9] | [91.2, 100.0] | [90.7, 100.0] |
| Day 36 | n (%) | 0 | 40 (100.0) | 38 (100.0) |
|  | [95% CI] | [0.0, 16.8] | [91.2, 100.0] | [90.7, 100.0] |
| Day 52 | n (%) | 0 | 40 (100.0) | 38 (100.0) |
|  | [95% CI] | [0.0, 16.8] | [91.2, 100.0] | [90.7, 100.0] |
| SARS-CoV-2 S1 NTD (AU/mL) |  |  |  |  |
| Day 22 | n (%) | 1 (5.0) | 40 (100.0) | 38 (100.0) |
|  | [95% CI] | [0.1, 24.9] | [91.2, 100.0] | [90.7, 100.0] |
| Day 36 | n (%) | 0 | 40 (100.0) | 38 (100.0) |
|  | [95% CI] | [0.0, 16.8] | [91.2, 100.0] | [90.7, 100.0] |
| Day 52 | n (%) | 0 | 40 (100.0) | 38 (100.0) |
|  | [95% CI] | [0.0, 16.8] | [91.2, 100.0] | [90.7, 100.0] |
